# Effectiveness of sexual health interventions for reducing risky sexual behaviors among people living with HIV: a network meta-analysis and systematic review

**DOI:** 10.1101/2025.09.13.25335688

**Authors:** Yixuan Li, Yanxiao Gao, Nancy Reynolds, Qiaoyue Lu, Ziqi Qin, Yuqiong Duan, Pingwu Wang, Tao Liu, Ling Jie Cheng, Wenru Wang, Honghong Wang, Xueling Xiao

## Abstract

**Objectives:** To assess and rank the effectiveness of different sexual health interventions designed to address risky sexual behaviors in PLWH.

**Methods:** We systematically searched studies published between Jan. 2010 and Dec. 2024. Randomized controlled trails (RCTs) were included if they recruited PLWH over 18 years old, reported original quantitative data on condomless behavior and multiple sexual partners across different groups. We calculated odds ratios (ORs) or standardized mean differences (SMDs) with 95% CIs to summarize the intervention effects.

**Results:** Our analysis included 34 RCTs with 12,967 participants across four intervention types. Goal-oriented intervention was most effective against the likelihood of engaging in condomless sex (OR: 0.54, 95%CI: 0.39 to 0.76), followed by safe sex skills training (OR: 0.65, 95%CI: 0.43 to 0.99). For reducing the number of condomless sex, goal-oriented intervention also ranked highest but showed no advantages compared to standard of care (SMD: −0.26, 95%CI: −-0.53 to 0.02). Six studies reported the data of multiple sexual partners among PLWH, no significant results were found.

**Conclusions:** Goal-oriented interventions should be prioritized in reducing condomless sex among PLWH. However, strategies to effectively address the issue of multiple sexual partners among PLWH remain unclear, underscoring the need for further high-quality evidence.

## Introduction

Risky sexual behaviors among people living with HIV (PLWH) remain a significant public health concern, as this population continue to be susceptible to co-infections despite the preventive benefits of antiretroviral therapy (ART) and the “Undetectable = Untransmittable” (U=U) paradigm (Tu et al., 2022; WHO, 2021). Evidence indicates that 27.9% of virally suppressed PLWH are newly diagnosed with other sexually transmitted infections (STIs) (Ge et al., 2023), and HIV superinfection continues to pose clinical challenges, with incidence rates reaching up to 7.7% annually (Redd et al., 2013), further exacerbates health burdens and complicates treatment regimens in this vulnerable population. While biomedical prevention strategies such as PrEP and PEP are effective for HIV-negative individuals, their population-level impact remains limited due to constrained accessibility and coverage, particularly in low-resource settings (Allan-Blitz & Mayer, 2024). In this context, advocating for consistent safe sex practices including encompassing protected sex and refraining from engaging in sexual activities with multiple partners, remain the most cost-effective and feasible approach. However, recent data indicated that the prevalence of risky sexual behaviors among PLWH remains alarmingly high, with rates ranging from 46% to 67% (Chen, 2019; Hamusonde et al., 2022; Worede et al., 2022), highlighting the urgent need to identify effective interventions for this population.

Numerous interventions have been developed to address the high prevalence of risky sexual behaviors among PLWH, yet this issue persists. Some conducted traditional health education intervention such as knowledge-based education, focusing on topics like sexual negotiation and condom use (Williams et al., 2012). Others emphasize mental health among vulnerable populations such as MSM living with HIV and newly-infected PLWH, supporting stress coping and promoting safer sex by alleviating symptoms of depression and anxiety (Brown et al., 2019; Pettifor et al., 2015). More are complex approaches, usually combined with other behavioral promotion, or interpersonal relationship improving (Holstad et al., 2012; Mashaphu et al., 2020). However, these interventions showed inconsistent effects, and some theoretically feasible methods are even discouraging (Xie et al., 2022).

Moreover, there are both inconsistency and heterogeneity in five published reviews summarizing the effectiveness of sexual health intervention for reducing risky sexual behaviors focus on PLWH. One meta-analysis confirmed a significant effect of “positive prevention” interventions in reducing the likelihood of condomless sex (Yin et al., 2014), while a 2021 meta also suggested that psychological interventions may increase the condom use in this population (Zhang et al., 2021). However, another review found these kind of interventions negatively impacted the reduction of sexual partners (Wang et al., 2013). Two additional reviews lacked meta-analyses: one U.S. review supported evidence-based interventions (Crepaz et al., 2014), while the other, summarizing behavioral interventions for older adult living with HIV, reported conflicting results among 6 studies (Illa et al., 2014). So, the effectiveness of existing interventions remains uncertain, with four of the five reviews were published over a decade ago, rendering them outdated. Advances in intervention contents, technology, and policies — such as eHealth interventions (Hirshfield et al., 2019), and “Undetectable = Untransmittable” (Bekker et al., 2023) campaign — may have further realigned intervention strategies. In response, we conducted a network meta-analysis aimed to assess and compare the effectiveness of different sexual health interventions in reducing risky sexual behaviors among PLWH.

## Methods

Our systematic review and network meta-analysis were conducted in accordance with Cochrane methodology for systematic reviews of effectiveness and followed the PRISMA guideline (Cumpston et al., 2019). The protocol registered in PROSPERO, and the registration number is CRD42023414692.

### Search strategies and selection criteria

We searched MEDLINE (PubMed), CINAHL (EBSCO), Excerpta Medica Database (Elsevier), Cochrane Central Register of Controlled Trials (Ovid), Web of Science (Clarivate) up to Jan 25, 2025. Searched grey literature database (https://opengrey.eu/) up to Jan 29, 2025, and searched two international trial registers (ClinicalTrials.gov, the World Health Organization’s International Clinical Trials Registry Platform) up to Jan 20, 2025. The search strategy was adapted for each included information source, and it was showed in Appendix-1.

We included RCTs that recruited people living with HIV aged 18 years or over, interventions that included sexual health promotion and reported condomless sex or multiple sexual partners. Studies involving both PLWH and individuals without HIV were also considered for inclusion if they reported original quantitative data specifically for the PLWH subgroup. We excluded studies that involve teenagers or children to ensure a focus on individuals who have attained legal and physiological maturity. We excluded studies published before 2010 since the introduction of “treatment as prevention” and PrEP have transformed strategies for reducing risky sexual behaviors among PLWH. If a single study publication contained multiple independent data samples, we presented them as separate reports.

### Study selection and data extraction

All searched studies were collected and imported into EndNote 20 software (Clarivate Analytics, Philadelphia, PA, USA), and duplicated references were removed. To better manage records and data throughout the review, references screening and data extraction were performed at Covidence platform (https://www.covidence.org/). Following a pilot test, studies were independently screened by two investigators (QL and QQ) for eligibility. Any disagreements were resolved through group discussion with YL, YD, XX or in consultant with other co-authors as needed. Data from eligible studies were extracted by three independent reviewers (PW, TL and QQ), they enlisted the aid of investigator (YL) or discuss with group members when disagreement arising. Data were collected from the included studies using a pre-structured data extraction form, which documented details including year of publication, the first author’s name, country, sample size, mean age, proportion of female participants, study setting, intervention format, deliverer and duration. Additionally, the “Ycasd” software was used to capture and scale data from graphical representations (Gross et al., 2014). We contacted the authors by email when required data was not available. The study was excluded if the author did not answer the email in three attempts within 7 days. The details of author connection and reason for study exclusion were shown in Appendix-4. In addition, One cluster RCT was included in this network meta-analysis and analyzed together with individual RCTs, following the best practice recommended by the Cochrane handbook (see Appendix 5).

### Assessment of risk of bias and confidence of evidence

Eligible studies were critically appraised by two independent reviewers (QL and QQ) using the revised Cochrane RoB 2.0 tool to assess risk of bias (Sterne et al., 2019). This tool evaluates bias in RCTs across five domains, including randomization, deviations from interventions, missing data, outcome measurement, and reporting. Overall bias is rated as low if all domains are low risk, some concerns if any domain has concerns, and high if one or more domains are high risk or multiple concerns compromise study validity. Conflicts were resolved by a third reviewer (YL) or group discussion. All studies, regardless of the results of their methodological quality, were undergo data extraction and synthesis. The strength of the body of evidence was assessed by the Grading of Recommendations Assessment, Development and Evaluation (GRADE) framework, the web application software the Confidence In Network Meta-Analysis (CINeMA) was used (Nikolakopoulou et al., 2020).

### Data synthesis and analysis

The primary outcome in this review was condomless sex and multiple sexual partners among PLWH including the proportion of participants who engaged in condomless sex /have two or more sexual partners and the mean number of condomless sex/sexual partners within a period of time. More details of data extraction and analyses for outcomes were described in Appendix-5.

The sexual health interventions were classified according to the description of elements, emphasis, primary goals, and mechanisms provided in each study. Additionally, it has been proved that different control conditions can lead to different estimations of effect size (Cuijpers & Cristea, 2016). Therefore, rather than aggregating all control groups into a single category, we meticulously classified them based on the specific descriptions outlined in each individual study. The characteristic and definition of interventions showed in Appendix-6.

We performed a standard pairwise meta-analysis using the “metan” package of Stata version 15.1. We applied DerSimonian and Laird random-effects model for pairwise meta-analyses. Standardized Mean Differences (SMDs) were used for continuous outcomes, such as the mean number of condomless sex acts, while Odds Ratios (ORs) were used for binary outcomes, such as the proportion of condomless sex, each reported with their respective 95% confidence intervals (CIs). Statistical heterogeneity was assessed using the I^2^ statistics, where an I^2^ > 50% indicates the presence of heterogeneity. In such cases, a random-effects model was used to calculate the pooled effect sizes. Otherwise, we would use a fixed-effect model. Besides, we assessed publication bias using the Egger’s test, and a p value > 0.05 indicated that there is no publication bias.

We performed a network meta-analysis (NMA) within a Bayesian framework. “gemtc” package in R software version 4.2.3 was used for computing a Markov chain Monte Carlo simulation. In NMA, we evaluated two models: a fixed-effect model and a random-effect model. We configured the model with four chains, performing 100,000 iterations and 50,000 annealing steps. The convergence of the model was assessed by visual inspection of 4 chains and the “Brooks-Gelman-Rubin” diagnostic method. We assessed the goodness of fit of each model to the data by calculating the posterior mean residual deviance. In addition, we used arm-based data; we estimated summary odds ratios (ORs) for binary outcome and the binomial likelihood was used, while standard mean differences (SMDs) for continuous outcome and the normal likelihood were used. Before conducted the network meta-analysis, we evaluated the transitivity considering clinical and methodological characteristics. Based on our knowledge, there is no standard way to analyze the transitivity across the different pairwise comparisons. To improve the reliability of results from network meta-analysis, we considered similar characteristics which would potentially associate with the result of effect size (Chaimani et al., 2017). Meanwhile, we asked for the opinions from two experts in the field of HIV/AIDS to check if there are other factors that should be considered. Finally, we investigated transitivity based on mean age, proportion of female, duration of intervention and sample size.

In network meta-analysis, the local and global inconsistency was evaluated by Stata, version 15.1 using “mvmeta” and “network” packages (Egger, 2008). The local inconsistency was checked by side-split approach and loop-specific approach, while the global inconsistency was checked by design-by-treatment method. For heterogeneity assessment, we assumed same heterogeneity for all treatment comparisons. We compared posterior distribution of the estimated heterogeneity variance (estimated tau^2^ for between-study heterogeneity) with its predictive distribution, defined as empirical distributions for tau^2^ provided by Turner et al for binary outcome and Rhodes et al for continuous outcome (Rhodes et al., 2015; Turner et al., 2015). We judged heterogeneity as very low when below the 25% quantile, as low when between 25% and 50% quantile, as moderate when between 50% and 75% and as high when above the 75% quantile (Bighelli et al., 2021).

The mean rank and surface under the cumulative ranking curve (SUCRA) were adopted to rank the treatment for each outcome. Publication bias was assessed by comparison-adjusted funnel plots, and Egger test employed to minimize the subjectivity of visual inspection. We conducted sensitivity analyses by excluding studies that focused on newly infected HIV-positive individuals, sample size ≤20, studies included HIV serodiscordant couples and studies with high risk of bias. The network diagrams were generated using the ‘mvmeta’ packages in Stata (version 15.1).

## Results

### Literature results

We identified 29178 results (29001 studies via database search and 177 studies from other resources), of which 6291 were duplicates and 6588 published before 2010. we did a full-text review of 133 of the 16299 studies, of which 94 were excluded (Figure 1). 34 studies including 41 samples of 12967 PLWH, were eligible in this review. Two studies reported both proportion and frequency of condomless sex, 21 studies reported proportion of condomless sex only, and 14 studies reported frequency of condomless sex only. Of these, 5 studies also reported the number of sexual partners, with another study reported the proportion of multiple sexual partners only. The Appendix-2 and 3 showed the list of included 34 RCTs and studies appears to meet the inclusion criteria but not included.

**Figure 1:**
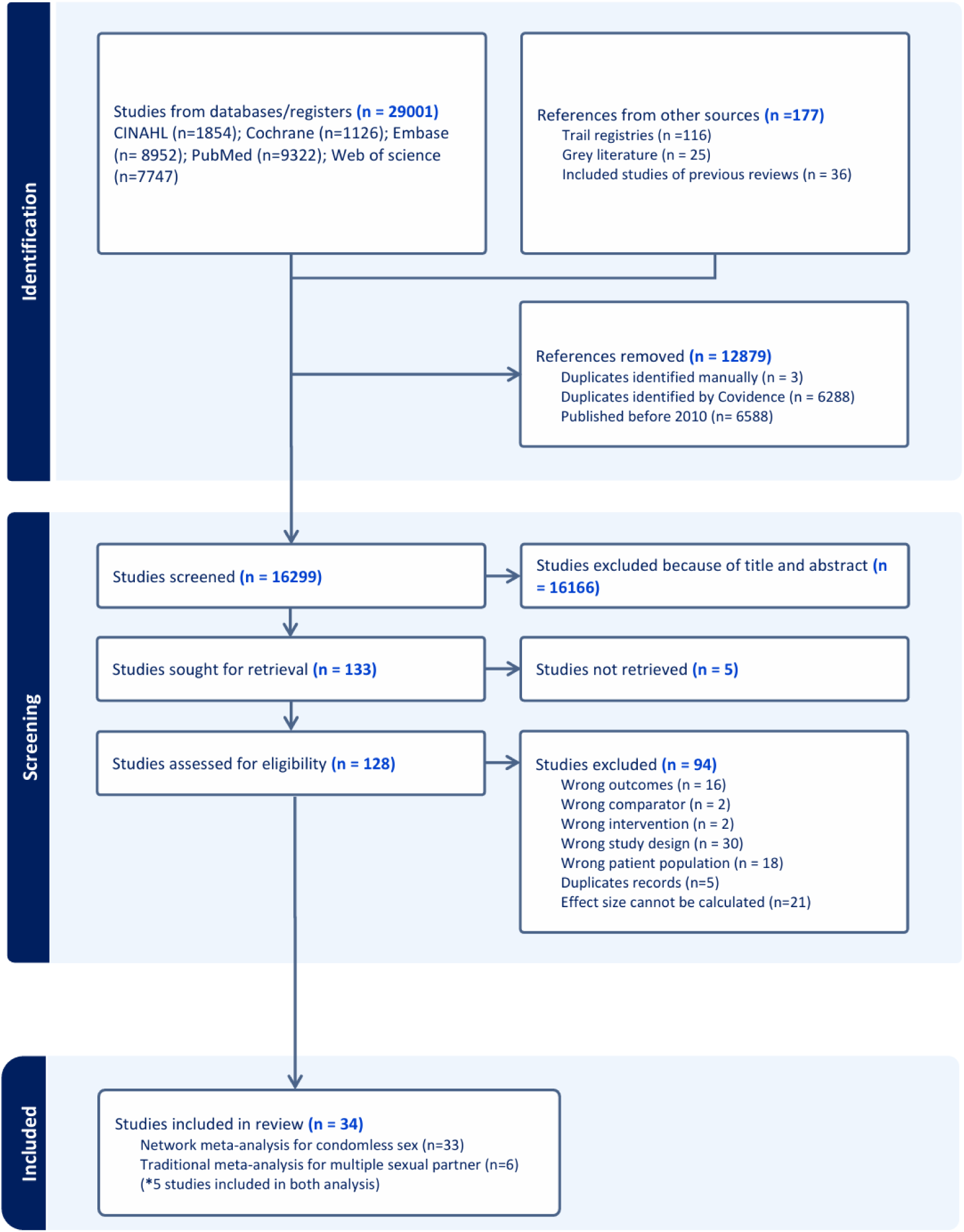
Study selection.

### Characteristics of included studies

Included studies were published from 2010 to 2022. The average number of participants per study was 393 (ranging from 21 to 2222) and the median intervention duration of the trials was 8 weeks (ranging from 1 to 48). Majority (n=21, 61.7%) of the studies recruited patients from North America, 11 (32.3%) from Africa, 1 (2.9%) from China, and 1(2.9%) from Russia. About 30% studies focused on HIV-positive MSM, and 5 trials recruited only woman living with HIV. 9 (26.4%) of 34 studies employed online interventions, with the majority of rest focused on group-based interventions delivered in person by trained health care workers (see Appendix-7).

### Traditional pairwise meta-analysis

The findings of the pairwise meta-analyses for condomless sex are shown in Appendix-10. For proportion outcome, goal-oriented intervention (GO) and education (Edu) presented statistically significantly advantage in reducing the likelihood of engaging in condomless sex compared with standard of care (GO for OR: 0.50, 95%CI: 0.34 to 0.74; Edu for OR: 0.63, 95%CI: 0.50 to 0.80). However, the other treatments did not show a statistical difference from the control conditions. For frequency outcome, we found a statistically significant difference in the two head-to-head comparisons: goal-oriented intervention VS standard of care (SMD: −0.12, 95%CI: −0.20 to −0.03) and safe sex skills training VS waiting list (SMD: −1.05, 95%CrI: −1.55 to −0.55). The Egger tests yielded no statistically significant results for any of the comparisons, indicating the absence of publication bias.

In addition, we conducted a meta-analysis of 6 studies reporting on multiple sexual partners (see Appendix-18). Among these, 3 studies measured the proportion of multiple sexual partners, while the other 3 assessed the mean number of partners. Compared to control groups, the intervention groups showed no significant advantages in reducing the likelihood of having two or more sexual partners (OR: 0.91, 95% CI: 0.70 to 1.17) or in decreasing the number of sexual partners (SMD: −0.03, 95% CI: - 0.61 to 0.55).

### Network meta-analysis

The network of eligible comparisons for condomless sex were showed in Figure 2. Four interventions and three control conditions formed two networks consisting of 7 nodes each, resulting in 21 network comparisons. 21 studies, including 8288 participants reported the proportion of PLWH engaged in condomless sex. Compared to standard of care, goal-oriented intervention showed a 46% reduction (OR: 0.54, 95%CI: 0.39 to 0.76) in the likelihood of engaging in condomless sex, and the safe sex skills training showed a 35% reduction (OR: 0.65, 95%CI: 0.43 to 0.99), see Figure 3A and Figure 4. Goal-oriented intervention also ranked highest among four interventions, with a SUCRA of 0.903, followed by safe sex skills training (0.727). But no statistically significant advantages over standard of care were found in decreasing the frequency of condomless sex in the past month (see Figure 3B), and the results of mixed comparison showed in Figure 4. All network meta-analysis results reported were obtained using the random-effects model, which provided a better fit compared to the fixed-effects model (see in Appendix-9).

**Figure 2.**
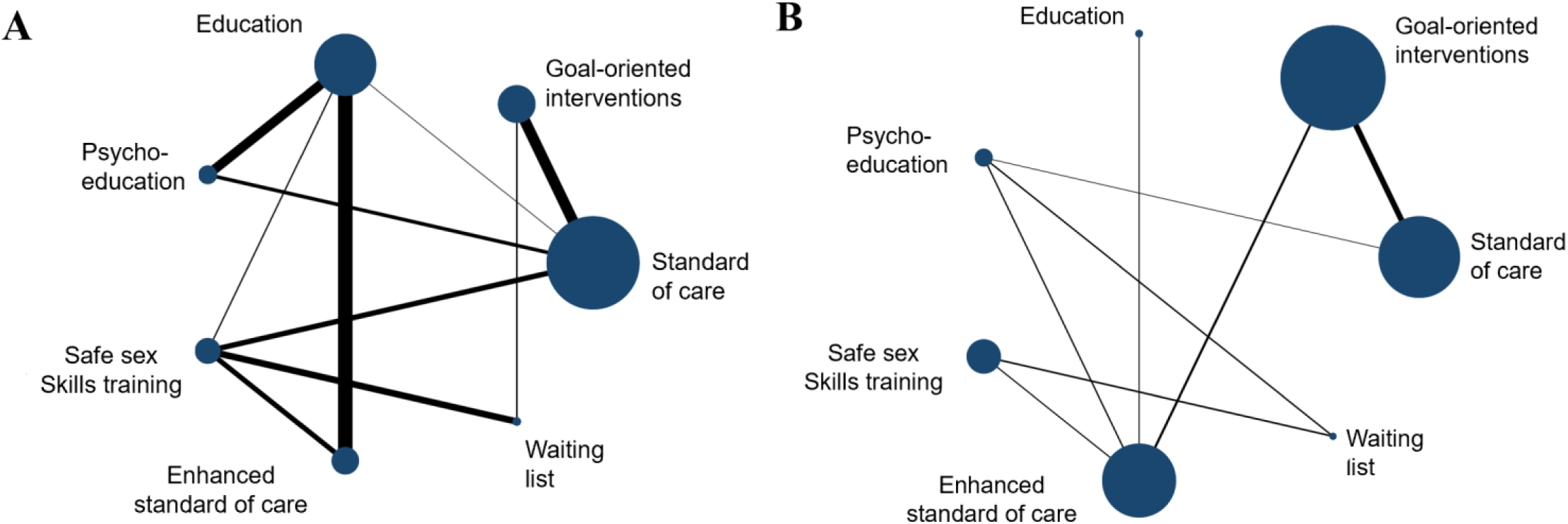
Network plot of eligible comparisons for proportion of condomless sex (A) and mean number of condomless sex in the past month (B) Each node represents an intervention, with the size of the node proportional to the number of participants randomly assigned (i.e., sample size), and the thickness of the lines proportional to the number of trials for each pair of treatments.

**Figure 3.**
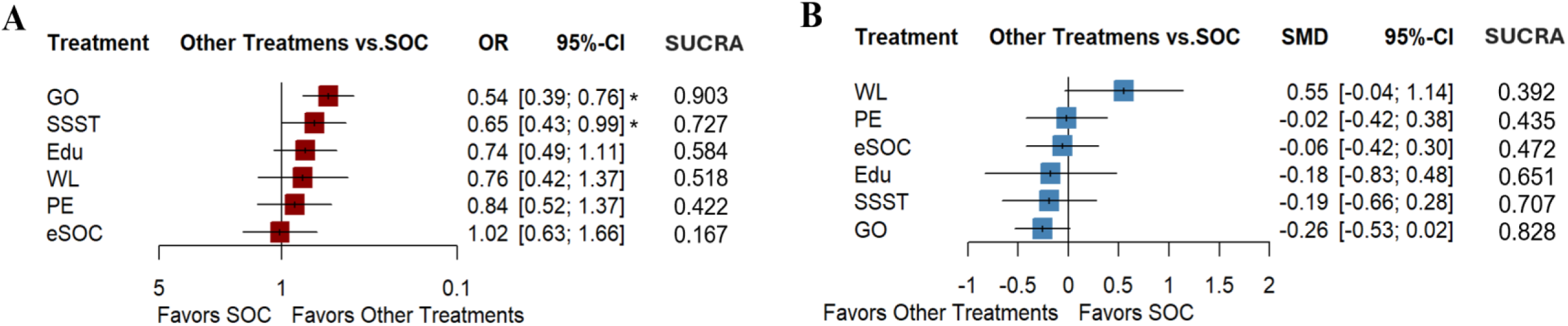
Forest plots of network meta-analysis for proportion of condomless sex (A) and mean number of condomless sex in the past month (B) OR, odds ratio; SMD, standardized mean differences; SUCRA, the surfaces under the cumulative ranking curve; 95% CI, 95% credible interval; *significant results. GO, goal-oriented intervention; WL, waiting list; SOC, standard of care; eSOC, enhanced standard of care; PE, psychoeducation; SSST, safe sex skills training; Edu, education.

**Figure 4.**
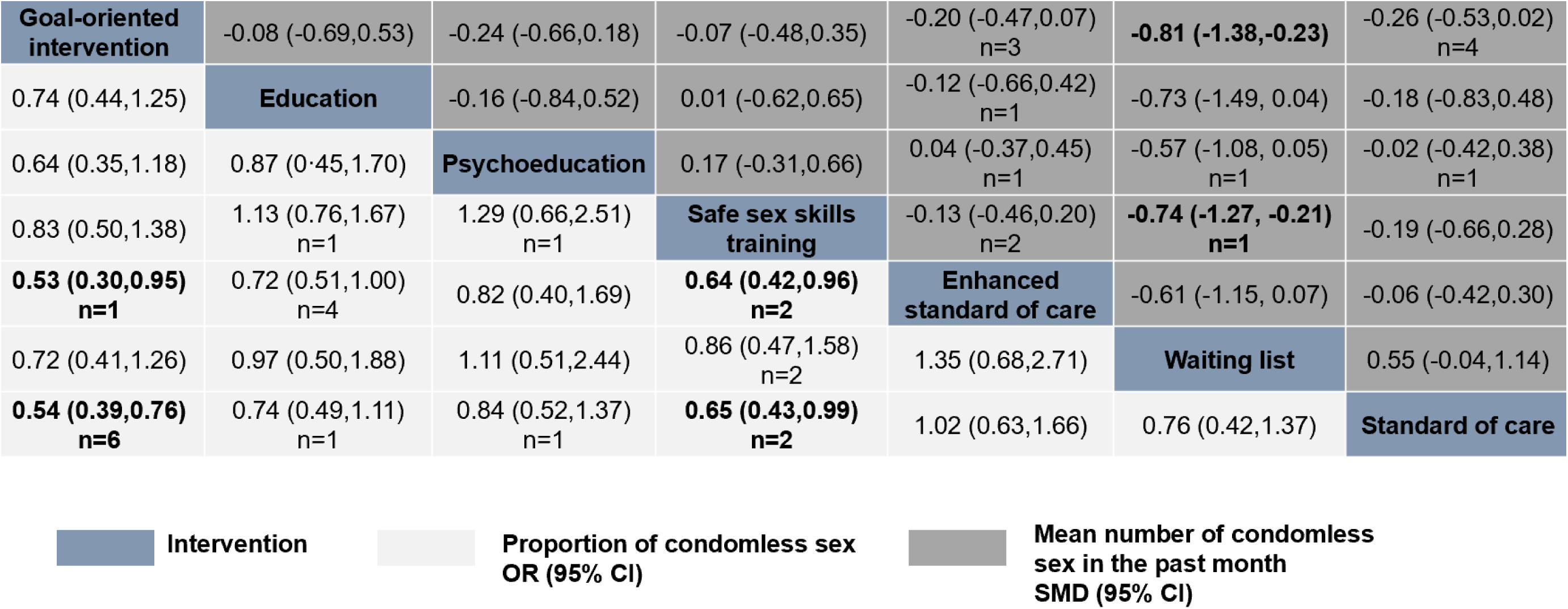
Results of mixed comparisons for proportion and frequency of the 4 sexual health interventions and 3 control conditions. Data are presented as ORs or SMDs (95%CI) for column-defined and row-defined treatments. The lower left triangle presents the results of proportion outcomes for ORs, and the upper right triangle presents the frequency outcomes for SMDs. ORs lower than 1 and SMDs lower than 0 favor the column-defining treatment. Significant results are presented in bold. N indicates the number of studies used for the comparison. OR, odds ratio; SMD, standardized mean differences; 95% CI, 95% credible interval.

We further conducted subgroup analyses for the goal-oriented intervention and safe sex skills training. For goal-oriented intervention, subgroup analyses of seven studies found no statistically significant change in the proportion outcome among PLWH across different country level of income, year of recruitment, intervention format or deliverer, but the confidence intervals included the possibility of no difference among MSM living with HIV. The high level of heterogeneity in studies was substantially lower, or absent, in study subgroups in which participants recruited after 2016, from high income country, intervention conducted online, and by self-help. (I^2^ range 0–26.5%; figures 5A). For safe sex skills training, we found no heterogeneity among seven studies that reported the proportion of condomless sex, but the estimates change to be no statistically difference both in all subgroups and the whole samples (figure 5B). Subgroup analyses for frequency outcome are presented in Appendix-12.

**Figure 5.**
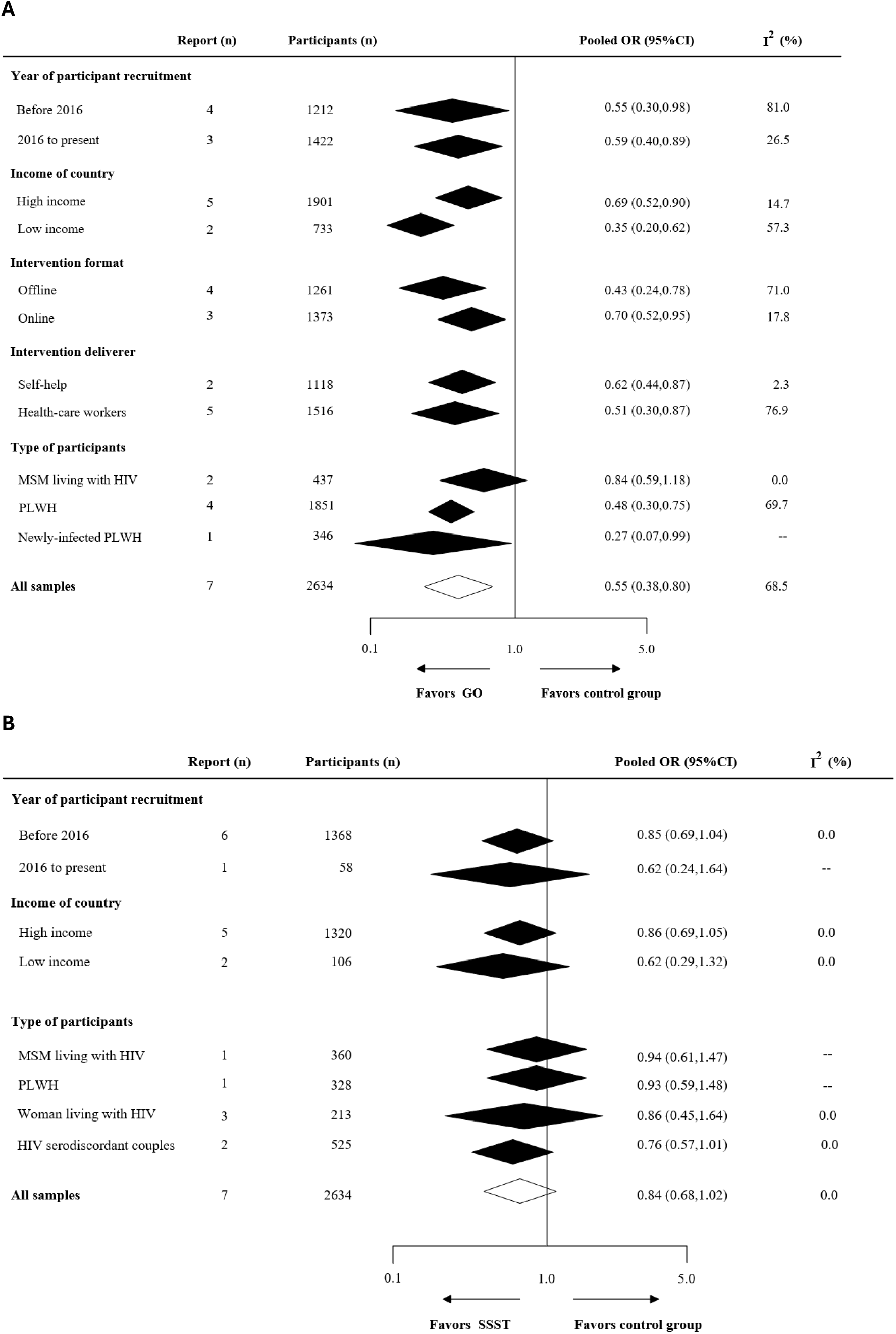
Subgroup meta-analysis of the effectiveness of goal-oriented intervention (A) and safe sex skills training (B) for reducing the prevalence of condomless sex. OR=odds ratio; PLWH=people living with HIV; MSM= men who have sex with men. GO=goal-oriented intervention; SSST=safe sex skills training.

### Transitivity, heterogeneity and inconsistency analysis

No definitive evidence was found to suggest violations of the transitivity assumption when evaluating potential effect modifiers across comparisons (see Appendix-13). Very low heterogeneity was observed in the proportion of condomless sex network (tau = 0.23, IQR 0.36–0.42), whereas low heterogeneity was noted in the frequency network (tau = 0.19, IQR 0.026–3.00) (see Appendix-14). Assessment of inconsistency showed in Appendix-15. The design-by-treatment test indicated no significant global inconsistency in both networks (proportion: p = 0. 3395; frequency: p = 0.3769). Locally, both of the proportion and frequency of condomless sex network demonstrated good consistency, as neither the loop-specific nor the side-splitting approach identified significant inconsistency (all p ≥ 0.05).

### Publication bias

The Supplementary Appendix-16 shows the results of comparison-adjusted funnel plot of both proportion and frequency outcomes, two plots are roughly symmetrical with egger test p = 0.304 for proportion and p = 0.224 for frequency. Thus, we considered no evidence of publication bias.

### Sensitivity analysis

In the sensitivity analysis, goal-oriented intervention still showed significant advantages over standard of care and ranked highest with the exception of studies include newly-infected patients, HIV serodiscordant couples, small sample studies, and studies rated as high risk of bias. However, for safe sex skills training, the confidence intervals included the possibility of no difference in majority of sensitivity analyses (see Appendix-17).

### Risk of bias and strength of evidence

Four studies were rated as low overall risk of bias, 11 studies had a high overall risk and with all remaining studies had a moderate overall risk (see Appendix-8). Confidence assessments using CINeMA ranged from low to moderate for most comparisons. Furthermore, we integrated the results of GRADE ratings in Figure 4, and the summarized results were shown in the Appendix-19.

## Discussion

This network systematic review and meta-analysis assessed and ranked the effectiveness of different sexual health interventions for reducing risky sexual behaviors among PLWH. Our analysis indicates that goal-oriented interventions are likely the most effective approach, showing significant advantages over control group in reducing both the likelihood and frequency of condomless sex. This effect remained consistent across various subgroups.

Overall, existing interventions emphasize setting measurable goals, enhancing knowledge, providing emotional support, and teaching practical skills to promote safer sexual practices. Despite the diverse focuses of existing interventions, a critical gap remains: none have explicitly integrated the U=U framework into their design or delivery (see Appendix-6). Accurate and context-sensitive communication of U=U is crucial: on the one hand, increasing numbers of national and international health agencies advocate for the widespread and accurate dissemination of the U=U message to ensure that PLWH are fully informed and can exercise their sexual health rights (Calabrese & Mayer, 2019). On the other hand, studies have shown that avoiding the delivery of U=U-related information is associated with persistent misconceptions about its meaning and implications (Rendina et al., 2020). Such misunderstandings may result in PLWH receiving inaccurate or incomplete information, which can lead to underestimation of the risks associated with engaging in condomless sex. Therefore, integrating U=U could enhance both the effectiveness and contextual relevance of sexual health interventions.

We found that both the goal-oriented intervention and safe sex skills training are more efficacious in reducing the likelihood and frequency of engaging in condomless sex than control conditions. However, in subgroup analyses, goal-oriented intervention consistently demonstrates significant superiority across different subgroups, whereas the benefits of safe sex skills training become less pronounced. This finding may indicate that the apparent effectiveness of safe sex skills training is largely influenced by indirect comparisons. Furthermore, there is no statistic difference between four sexual health interventions for both proportion and frequency outcomes. Possible explanations could be the limit number of included trials conducting education, psychoeducation and safe sex skills training interventions (most of direct comparison from 1 to 2 trials), as well as the fact that the comparative results for intervention conditions are mostly derived from indirect comparisons.

The superior effect of goal-oriented interventions aligns with prior evidence showing their effectiveness in promoting health behavior change (Bailey, 2019). Sexual behaviors among PLWH involve a complex decision-making process characterized by unpredictability, instability, and influenced by multiple factors. Goal-oriented interventions offer unique advantages in addressing these challenges, primarily by implying two key components: goal setting and action planning. Setting clear and specific goals tailored to the individual’s circumstances, such as aiming to reduce the number of sexual partners over the next six months or practicing refusal skills to avoid unprotected sex in high-risk situations every week, can effectively stimulate intrinsic motivation (Hart et al., 2021; Kahler et al., 2018), improve adherence to action plans, thereby supporting behavior change (Shilts et al., 2013; Wilson & Brookfield, 2009). This benefit may be attributed to the goal-setting process, where individuals gain a clearer understanding of the risks they face and increase their acceptance of specific behavioral changes needed. Besides, detailed action plans help reduce the challenges associated with behavior change. These plans may include steps such as obtaining and using condoms correctly, and negotiating safer practices with partners.

However, despite these benefits, goal-oriented intervention maybe challenges in effectively managing condomless sex among MSM living with HIV. One potential reason for this limitation is the compounded self-stigma associated with both HIV status and MSM identity, which goal-oriented interventions may not fully address (van Bilsen et al., 2020). Additionally, the dynamics of sexual relationships and risk behaviors among MSM community are often complex and changeable (Reisner et al., 2016). So, sexual health intervention among MSM living with HIV may require more personalized and real-time monitoring approaches.

According to our SUCRA results for both proportion and frequency outcomes, education and safe sex skills training might rank second and third among the four sexual health interventions. Safe sex skills training showed significant advantages over the control conditions for reducing both the proportion and frequency of the condomless sex and it was most likely to rank second out of the four sexual health interventions. But the statistical estimates were not stable in subgroup and sensitivity analysis. A similar pattern was observed with educational interventions, which remain widely used and are incorporated into sexual health service guidelines for people living with HIV (Ford et al., 2021). However, these approaches may lack of consideration for the unique needs of different individuals. Moreover, previous reviews have highlighted that tailored interventions are more effective than non-tailored interventions (Kim et al., 2021), offering greater sensitivity in addressing the needs of vulnerable and socially marginalized populations (Meiksin et al., 2021). Certainly, the potential reason for the unstable and negative results may be associated with the limited included studies. Therefore, more large-sample and high-quality RCTs should be done to identify the effectiveness of educational and skills training interventions.

Interestingly, psychoeducation had poor SUCRA values in both networks, even ranking below some control conditions. This finding contrasts with previous review indicating that psychological interventions positively affect the reduction of sexual risk among PLWH (Zhang et al., 2021). This discrepancy may stem from Zhang’s broader definition of psychological interventions, whereas the psychoeducation interventions in our analysis specifically focused on emotional support and coping strategies. Furthermore, some studies have reported no association between depression and risky sexual behaviors (Tsai et al., 2013), with evidence suggesting that PLWH experiencing severe depression may be less inclined to engage in risky sexual behaviors due to the preponderance of debilitating neurovegetative symptoms (O’Cleirigh et al., 2013). Thus, psychoeducation may not be a universally effective intervention, individualized assessments are likely necessary before its application.

To our knowledge, this is the first network systematic review and meta-analysis to investigate the effectiveness of different sexual health interventions for reducing risky sexual behaviors among PLWH, offering practical insights for HIV healthcare providers. However, our review has some limitations. First, nearly one-third of included studies were rated as high risk of bias, indicating that our results should be interpreted with caution. However, there may be uncontrolled factors such as personal motivation, cultural background, social support, and impossible double-blinded in sexual health intervention. Second, the limited number of studies that reported multiple sexual partners prevented us from conducting a network meta-analysis. Third, differences in populations, such as newly-infected individuals and HIV serodiscordant couples, may affect the reliability of the evidence. Despite this, the subgroup analyses and sensitivity analyses did not provide substantial evidence suggesting violations of the transitivity and consistency across different populations. Finally, we unable to conduct subgroup analysis by follow-up time as only two studies followed participants for more than one year, making it difficult to assess the long-term sustainability of intervention effects.

Based on our findings, we recommend that HIV service organizations and healthcare providers prioritize goal-oriented interventions, particularly those incorporating goal-setting and planning, when designing strategies to reduce risky sexual behaviors among PLWH. However, there are significant gaps in the evidence: no interventions address the U=U (“Undetectable = Untransmittable”) message, potentially leaving patients unaware of how to engage in safer sexual practices. Additionally, there is a lack of studies targeting the reduction of multiple sexual partners, and it remains uncertain whether intervention effectiveness varies across different populations and follow-up durations or if other potentially effective interventions exist. Therefore, further research with robust study designs, extended follow-up periods, targeted evaluation of interventions aimed at reducing the number of sexual partners, and direct comparisons of alternative strategies is needed. Equally important is ensuring the precise communication of U=U principles within health care delivery.

## Conclusions

The results of this review have important implications for the prevention of further co-infection among PLWH, including HIV superinfection and other STI. We found that goal-oriented interventions, more specifically, personalized goal setting and planning, are most likely to be the optimal approach in reducing the likelihood of engaging in condomless sex among PLWH. Given the high prevalence of risky sexual behaviors, limited access to biomedical prevention, and residual risks despite viral suppression, prioritizing goal-oriented interventions identified as the most effective in this review may offer a feasible and cost-effective strategy to reduce co-infections and broader health risks among PLWH.

## Data Availability

This systematic review and network meta-analysis did not involve the collection of primary data and thus did not require ethical approval. All data were obtained from publicly available sources.

## Author Contributions

The study was designed by YL and XX, with contributions from YG, WW, HW, NRR and CLJ. YL and QL set up the search term, with help from CLJ and HW. YL, ZQ and QL completed the study selection, data extraction and quality assessment, YG, XX, HW, and WW joined in the group discussion when conflict exist. ZQ, PW, TL and YD acquired possible trials from relevant reviews by screening the reference list and contacting the authors. ZQ, PW, TL and YD verified the data, QL and YL did the statistical analysis, with the help of CLJ and YG. YL, YG and XX draft the manuscript, WW, HW, NRR and CLJ critically reviewed and editing the manuscript. All authors had complete access to the study data and take responsibility for its submission for publication.

