## Supplementary material for "Effectiveness of sexual health interventions for reducing risky sexual behaviors among people living with HIV: a network meta-analysis and systematic review": Supplimental files

### Supplementary Appendix

#### *Appendix-1: Search strategy*

Before undertaking this systematic review and meta-analysis, we did a preliminary search. The search database MEDLINE (PubMed), Excerpta Medica Database (Elsevier), JBI systematic review register、the Cochrane Central Register of Controlled Trials, PROSPERO, and The Clinical Trials were performed to check whether there were existing or ongoing review protocol. The process of systematic search are follows: Firstly, an initial limited search of PubMed was undertaken to identify articles on the topic. Secondly, the text words contained in the titles, abstracts or keywords of relevant articles, and the index terms used to describe the articles were used to develop a primary search strategy. Thirdly, a full search strategy was adapted from previous related reviews and improved by experts in HIV/AIDS and behavioral intervention fields. Additionally, we checked the references of related reviews [1,2] in order to retrieve relevant articles which have not been captured in database searches. The full search strategy of each database are follows:

MEDLINE (PubMed)

1. "HIV Infections"[MeSH Major Topic:noexp]/ 202393
2. "Acquired Immunodeficiency Syndrome"[MeSH Major Topic:noexp]/ 63493
3. "HIV"[MeSH Major Topic:noexp]/ 12269
4. (((((HIV infect*[Title/Abstract]) OR (HIV positiv*[Title/Abstract])) OR (living with HIV[Title/Abstract])) OR (Acquired Immune Deficiency Syndrome[Title/Abstract])) OR (HIV NEAR/4 diagnos*[Title/Abstract])) OR (AIDs[Title/Abstract])/ 295819
5. 1 or 2 or 3 or 4/ 392047
6. Sexual behavior [MeSH Major Topic:noexp]/ 34937
7. ((((((Sexual Activit*[Title/Abstract]) OR (Sexual Behavio*[Title/Abstract])) OR (Oral Sex[Title/Abstract])) OR (anal intercourse[Title/Abstract])) OR (vaginal intercourse [Title/Abstract])) OR (Anal Sex[Title/Abstract])) OR (vaginal sex[Title/Abstract])/ 47316
8. ((((((((risk-taking[Title/Abstract]) OR (risk behavio*[Title/Abstract])) OR (risk practice[Title/Abstract])) OR (risk activit*[Title/Abstract])) OR (condom*[Title/Abstract])) OR (unsafe sex*[Title/Abstract])) OR (unprotected

sex*[Title/Abstract])) OR (sexual risk[Title/Abstract])) OR (sexual partner*[Title/Abstract])/ 66103

9. 5 or 6 or 7 or 8/ 476,314

10. ((((((((Intervention[Title/Abstract]) OR (therap*[Title/Abstract])) OR (reduc*[Title/Abstract])) OR (train*[Title/Abstract])) OR (treatment[Title/Abstract])) OR (promot*[Title/Abstract])) OR (support*[Title/Abstract])) OR (strateg*[Title/Abstract])) OR (program[Title/Abstract])) OR (counsel*[Title/Abstract])/ 14,291,740

11. (((((((((skill[Title/Abstract]) OR (deliver*[Title/Abstract])) OR (service*[Title/Abstract])) OR (prevention[Title/Abstract])) OR (experiment*[Title/Abstract])) OR (coping[Title/Abstract])) OR (approach*[Title/Abstract])) OR (psychotherap*[Title/Abstract])) OR (psychoeducation*[Title/Abstract])) OR (cognitive*[Title/Abstract])) OR (motivation*[Title/Abstract])/ 7,373,925

12. 10 or 11/ 17,630,610

13. randomized controlled trial[pt]/ 622,284

14. controlled clinical trial[pt]/ 712,967

15. (((((random*[Title/Abstract]) OR (placebo[Title/Abstract])) OR (group*[Title/Abstract])) OR (trail*[Title/Abstract])) OR (RCT[Title/Abstract])) OR (control* [Title/Abstract])/ 6,222,477

16. 13 or 14 or 15/ 6,332,947

17. 5 and 9 and 12 and 16/ 9322

Web of Science (Clarivate)

1. (((((AB=("hiv infect*")) OR AB=("hiv positiv*")) OR AB=("living with hiv")) OR AB=("Acquired Immune Deficiency Syndrome")) OR AB=("HIV NEAR/4 diagnos*")) OR AB=("AIDs")/ 241,716

2. (((((TI=("hiv infect*")) OR TI=("hiv positiv*")) OR TI=("living with hiv")) OR TI=("Acquired Immune Deficiency Syndrome")) OR TI=("HIV NEAR/4 diagnos*")) OR TI=(AIDs)/ 254,882

3. 1 or 2/ 421,774

4. ((((((((((((((TI=("Sexual Activit*")) OR TI=("Sexual Behavio*")) OR TI=("Oral Sex")) OR TI=("anal intercourse")) OR TI=("vaginal intercourse")) OR TI=("Anal Sex")) OR TI=("vaginal sex")) OR TI=("risk-taking")) OR TI=("risk behavio*")) OR TI=("risk practice")) OR TI=("risk activit*")) OR TI=("condom*")) OR TI=("unsafe sex*")) OR TI=("unprotected sex*")) OR TI=("sexual risk")) OR TI=("sexual partner*")/ 39208

5. ((((((((((((((AB=("Sexual Activit*")) OR AB=("Sexual Behavio*")) OR AB=("Oral Sex")) OR AB=("anal intercourse")) OR AB=("vaginal intercourse")) OR AB=("Anal Sex")) OR AB=("vaginal sex")) OR AB=("risk-taking")) OR AB=("risk behavio*")) OR AB=("risk practice")) OR AB=("risk activit*")) OR AB=("condom*")) OR AB=("unsafe sex*")) OR AB=("unprotected sex*")) OR AB=("sexual risk")) OR AB=("sexual partner*")/ 102115

6. 4 or 5/ 115,871

7. (((((((SO=("randomized controlled trial")) OR SO=("controlled clinical trial")) OR TI=(random*)) OR TI=(placebo)) OR TI=(group*)) OR TI=(trail*)) OR TI=(RCT)) OR TI=(controlled)/ 2868793

8. (((((AB=(random*)) OR AB=(placebo)) OR AB=(group*)) OR AB=(trail*)) OR AB=(RCT)) OR AB=(controlled)/ 13678211

9. 7 or 8/ 14,764,517

10. ((((((((((((((((((((TI=(Intervention)) OR TI=(therap*)) OR TI=(reduc*)) OR TI=(train*)) OR TI=(treatment)) OR TI=(promot*)) OR TI=(support*)) OR TI=(skill)) OR TI=(strateg*)) OR TI=(deliver*)) OR TI=(service*)) OR TI=(prevention)) OR TI=(cbt)) OR TI=(program)) OR TI=(counsel*)) OR TI=(experiment*)) OR TI=(coping)) OR TI=(approach*)) OR TI=(psychotherap*))

OR TI=(psychoeducation*)) OR TI=(cognitive*)) OR TI=(motivation*)/ 9544176

11. ((((((((((((((((((((AB=(Intervention)) OR AB=(therap*)) OR AB=(reduc*)) OR AB=(train*)) OR AB=(treatment)) OR AB=(promot*)) OR AB=(support*)) OR AB=(skill)) OR AB=(strateg*)) OR AB=(deliver*)) OR AB=(service*)) OR AB=(prevention)) OR AB=(cbt)) OR AB=(program)) OR AB=(counsel*)) OR AB=(experiment*)) OR AB=(coping)) OR AB=(approach*)) OR AB=(psychotherap*)) OR AB=(psychoeducation*)) OR AB=(cognitive*)) OR AB=(motivation*)/ 30404802

12. 10 or 11/ 36380369

13. 3 and 6 and 9 and 12/ 7747

Excerpta Medica Database (Elsevier)

1. 'human immunodeficiency virus infection'/mj 194698

2. 'hiv infect*':ab,ti OR 'hiv positiv*':ab,ti OR 'living with hiv':ab,ti OR 'acquired immune deficiency syndrome':ab,ti OR ((hiv NEAR/4 diagnos*):ab,ti) OR aids:ab,ti 364251

3. 1 or 2 420581

4. 'sexual behavior'/mj 15890

5. 'sexual activit*':ab,ti OR 'sexual behavio*':ab,ti OR 'oral sex':ab,ti OR 'anal intercourse':ab,ti OR 'vaginal intercourse':ab,ti OR 'anal sex':ab,ti OR 'vaginal sex':ab,ti OR 'risk taking':ab,ti OR 'risk behavio*':ab,ti OR 'risk practice':ab,ti OR 'risk activit*':ab,ti OR condom*:ab,ti OR 'unsafe sex*':ab,ti OR 'unprotected sex*':ab,ti OR 'sexual risk':ab,ti OR 'sexual partner*':ab,ti 128673

6. 4 or 5 138426

7. 'randomized controlled trial':it OR 'controlled clinical trial':it OR random*:ab,ti OR placebo:ab,ti OR group*:ab,ti OR trail*:ab,ti OR rct:ab,ti OR controlled:ab,ti 7592198

8. intervention:ab,ti OR therap*:ab,it OR reduc*:ab,ti OR train*:ab,ti OR treatment:ab,ti OR promot*:ab,ti OR support*:ab,ti OR skill:ab,ti OR strateg*:ab,ti

OR deliver*:ab,ti OR service*:ab,ti OR prevention:ab,ti OR cbt:ab,ti OR program:ab,ti OR counsel*:ab,ti OR experiment*:ab,ti OR coping:ab,ti OR approach*:ab,ti OR psychotherap*:ab,ti OR psychoeducation*:ab,ti OR cognitive*:ab,ti OR motivation*:ab,ti 18637357

9. 3 and 6 and 7 and 8 8952

Cochrane Central Register of Controlled Trials (Ovid)

1. MeSH descriptor [HIV Infections] this term only 11921
2. "HIV Infect*":ti,ab OR "aids":ti,ab OR "living with HIV":ti,ab OR "HIV NEAR/4 diagnos*":ti,ab OR "HIV positiv*":ti,ab OR "AIDS":ti,ab OR "acquired immune deficiency syndrome":ti,ab 12058
3. 1 or 2 21568
4. MeSH descriptor [Sexual behavior] this term only 3195
5. "Sexual Activit*":ti,ab OR "Sexual Behavio*":ti,ab OR "Oral Sex":ti,ab OR "anal intercourse":ti,ab OR "vaginal intercourse":ti,ab OR "Anal Sex":ti,ab OR "vaginal sex":ti,ab OR "risk-taking":ti,ab OR "risk behavio*":ti,ab OR "risk practice":ti,ab OR "risk activit*":ti,ab OR "condom*":ti,ab OR "unsafe sex*":ti,ab OR "unprotected sex*":ti,ab OR "sexual risk":ti,ab OR "sexual partner*":ti,ab 5312
6. 4 or 5 6420
7. "randomized controlled trial":pt OR "controlled clinical trial":pt OR "random*":ti,ab OR "placebo":ti,ab OR "group*":ti,ab OR "trail*":ti,ab OR "RCT":ti,ab OR "controlled":ti,ab 1145293
8. "Intervention":ti,ab OR "therap*":ti,ab OR "reduc*":ti,ab OR "train*":ti,ab OR "treatment":ti,ab OR "promot*":ti,ab OR "support*":ti,ab OR "skill":ti,ab OR "strateg*":ti,ab OR "deliver*":ti,ab OR "service*":ti,ab OR "prevention":ti,ab OR "CBT":ti,ab OR"program":ti,ab OR "counsel*":ti,ab OR "experiment*":ti,ab OR "coping":ti,ab OR "approach*":ti,ab OR "psychotherap*":ti,ab OR

"psychoeducation*":ti,ab OR "cognitive*":ti,ab OR "motivation*":ti,ab 1193251

9. 3 and 6 and 7 and 8 1126

CINAHL (EBSCO)

1. AB "HIV infect*" OR AB "HIV positiv*" OR AB "living with HIV" OR AB "Acquired Immune Deficiency Syndrome" OR AB "HIV W4 diagnos*" OR AB "AIDs" 55772
2. TI "HIV infect*" OR TI "hiv positiv*" OR TI "living with HIV" OR TI "Acquired Immune Deficiency Syndrome" OR TI "HIV M4 diagnos*" OR TI "AIDs" 43047
3. 1 or 2 78409
4. TI "Sexual Activit*" OR TI "Sexual Behavio*" OR TI "Oral Sex" OR TI "anal intercourse" OR TI "vaginal intercourse" OR TI "Anal Sex" OR TI "vaginal sex" OR TI "risk-taking" OR TI "risk behavio*" OR TI "risk practice" OR TI "risk activit*" OR TI "condom*" OR TI "unsafe sex*" OR TI "unprotected sex*" OR TI "sexual risk" OR TI "sexual partner*" 12256
5. AB"Sexual Activit*" OR AB "Sexual Behavio*" OR AB "Oral Sex" OR AB "anal intercourse" OR AB "vaginal intercourse" OR AB "Anal Sex" OR AB "vaginal sex" OR AB "risk-taking" OR AB "risk behavio*" OR AB "risk practice" OR AB "risk activit*" OR AB "condom*" OR AB="unsafe sex*" OR AB "unprotected sex*" OR AB "sexual risk" OR AB "sexual partner*" 29368
6. 4 or 5 33799
7. TI "random*" OR TI "placebo" OR TI "group*" OR TI "trail*" OR TI "RCT" OR TI "controlled" 238973
8. AB "random*" OR AB "placebo" OR AB "group*" OR AB "trail*" OR AB "RCT" OR AB "controlled" 1190809
9. 7 or 8 1284494

10. TI "Intervention" OR TI "therap*" OR TI "reduc*" OR TI "train*" OR TI "treatment" OR TI "promot*" OR TI "support*" OR TI "skill" OR TI "strateg*" OR TI "deliver*" OR TI "service*" OR TI "prevention" OR TI "CBT" OR TI "program" OR TI "counsel*" OR TI "experiment*" OR TI "coping" OR TI "approach*" OR TI "psychotherap*" OR TI "psychoeducation*" OR TI "cognitive*" OR TI "motivation*" 1428179

11. AB "Intervention" OR AB "therap*" OR AB "reduc*" OR AB "train*" OR AB "treatment" OR AB "promot*" OR AB "support*" OR AB "skill" OR AB "strateg*" OR AB "deliver*" OR AB "service*" OR AB "prevention" OR AB "CBT" OR AB "program" OR AB "counsel*" OR AB "experiment*" OR AB "coping" OR AB "approach*" OR AB "psychotherap*" OR AB "psychoeducation*" OR AB "cognitive*" OR AB "motivation*" 2692612

12. 10 or 11 3383168

13. 3 and 6 and 9 and 12 1854

#### *Appendix-2: List of included 34 randomized controlled trials*

**1. Bachanas 2016**

Bachanas, Pamela et al. "Delivering Prevention Interventions to People Living with HIV in Clinical Care Settings: Results of a Cluster Randomized Trial in Kenya, Namibia, and Tanzania." AIDS and behavior vol. 20,9 (2016): 2110-8. doi:10.1007/s10461-016-1349-2

**2. Brown 2019**

Brown, J. L., Vanable, P. A., Bostwick, R. A., & Carey, M. P. (2019). A Pilot Intervention Trial to Promote Sexual Health and Stress Management Among HIV-Infected Men Who Have Sex with Men. AIDS Behav, 23(1), 48-59. https://doi.org/10.1007/s10461-018-2234-y

**3. Cruess 2018**

Cruess, D. G., Burnham, K. E., Finitsis, D. J., Goshe, B. M., Strainge, L., Kalichman, M., Grebler, T., Cherry, C., & Kalichman, S. C. (2018). A Randomized Clinical Trial of a Brief Internet-based Group Intervention to Reduce Sexual Transmission Risk Behavior Among HIV-Positive Gay and Bisexual Men. Ann Behav Med, 52(2), 116-129. https://doi.org/10.1093/abm/kax031

**4. Dévieux 2022**

Dévieux, J. G., Rosenberg, R., Jean-Gilles, M., Villalba, K., Attonito, J., Lerner, B., Saxena, A., & Stein, J. (2022). Effectiveness of a Cognitive Behavioral Randomized Controlled Trial for People Living with HIV Who are Heavy Drinkers: The Holistic Health Recovery Program (HHRP) Trial in Miami. J Clin Psychol Med Settings, 29(3), 498-508. https://doi.org/10.1007/s10880-022-09878-5

**5. Echenique 2013**

Echenique, M., Illa, L., Saint-Jean, G., Avellaneda, V. B., Sanchez-Martinez, M., & Eisdorfer, C. (2013). Impact of a secondary prevention intervention among HIV-positive older women. AIDS Care, 25(4), 443-446. https://doi.org/10.1080/09540121.2012.712666

**6. Fisher 2014**

Fisher, J. D., Cornman, D. H., Shuper, P. A., Christie, S., Pillay, S., Macdonald, S., Ngcobo, N., Amico, K. R., Lalloo, U., Friedland, G., & Fisher, W. A. (2014). HIV prevention counseling intervention delivered during routine clinical care reduces HIV risk behavior in HIV-infected South Africans receiving antiretroviral therapy: the Izindlela Zokuphila/Options for Health randomized trial. J Acquir Immune Defic Syndr, 67(5), 499-507. https://doi.org/10.1097/qai.0000000000000348

**7. Golin 2013**

Golin, C. E., Earp, J. A., Grodensky, C. A., Patel, S. N., Suchindran, C., Parikh, M., Kalichman, S., Patterson, K., Swygard, H., Quinlivan, E. B., Amola, K., Chariyeva, Z., & Groves, J. (2013). Longitudinal effects of SafeTalk, a motivational interviewing-based program to improve safer sex practices among people living with HIV/AIDS. AIDS Behav, 16(5), 1182-1191. https://doi.org/10.1007/s10461-011-0025-9

**8. Hart 2021**

Hart, T. A., Noor, S. W., Skakoon-Sparling, S., Lazkani, S. N., Gardner, S., Leahy, B., Maxwell, J., Julien, R., Simpson, S., Steinberg, M., & Adam, B. D. (2021). GPS: A Randomized Controlled Trial of Sexual Health Counseling for Gay and Bisexual Men Living With HIV. Behav Ther, 52(1), 1-14. https://doi.org/10.1016/j.beth.2020.04.005

**9. Kahler 2018**

Kahler, C. W., Pantalone, D. W., Mastroleo, N. R., Liu, T., Bove, G., Ramratnam, B., Monti, P. M., & Mayer, K. H. (2018). Motivational interviewing with personalized feedback to reduce alcohol use in HIV-infected men who have sex with men: A randomized controlled trial. J Consult Clin Psychol, 86(8), 645-656. https://doi.org/10.1037/ccp0000322

**10. Kalichman 2018**

Kalichman, S. C., Cherry, C., Kalichman, M. O., Eaton, L. A., Kohler, J. J., Montero, C., & Schinazi, R. F. (2018). Mobile Health Intervention to Reduce HIV Transmission: A Randomized Trial of Behaviorally Enhanced HIV Treatment as Prevention (B-TasP). J Acquir Immune Defic Syndr, 78(1), 34-42. https://doi.org/10.1097/qai.0000000000001637

**11. Klein 2013**

Klein, C. H., Lomonaco, C. G., Pavlescak, R., & Card, J. J. (2013). WiLLOW: reaching HIV-positive African-American women through a computer-delivered intervention. AIDS Behav, 17(9), 3013-3023. https://doi.org/10.1007/s10461-013-0479-z

**12. Kurth 2014**

Kurth, A. E., Spielberg, F., Cleland, C. M., Lambdin, B., Bangsberg, D. R., Frick, P. A., Severynen, A. O., Clausen, M., Norman, R. G., Lockhart, D., Simoni, J. M., & Holmes, K. K. (2014). Computerized counseling reduces HIV-1 viral load and sexual transmission risk: findings from a randomized controlled trial. J Acquir Immune Defic Syndr, 65(5), 611-620. https://doi.org/10.1097/qai.0000000000000100

**13. Liu 2018**

Liu, Y., Vermund, S. H., Ruan, Y., Liu, H., Rivet Amico, K., Simoni, J. M., Shepherd, B. E., Shao, Y., & Qian, H. Z. (2018). Peer counselling versus standard-of-care on reducing high-risk behaviours among newly diagnosed HIV-positive men who have sex with men in Beijing, China: a randomized intervention study. J Int AIDS Soc, 21(2). https://doi.org/10.1002/jia2.25079

**14. Milam 2016**

Milam, Joel et al. “Randomized Controlled Trial of an Internet Application to Reduce HIV Transmission Behavior Among HIV Infected Men Who have Sex with Men.” AIDS and behavior vol. 20,6 (2016): 1173-1181.

**15. Holstad 2012**

Holstad, M. M., Essien, J. E., Ekong, E., Higgins, M., Teplinskiy, I., & Adewuyi, M. F. (2012). Motivational groups support adherence to antiretroviral therapy and use of risk reduction behaviors in HIV positive Nigerian women: a pilot study. Afr J Reprod Health, 16(3), 14-27.

**16. Marhefka 2014**

Marhefka, S. L., Buhi, E. R., Baldwin, J., Chen, H., Johnson, A., Lynn, V., & Glueckauf, R. (2014). Effectiveness of healthy relationships video-group-A videoconferencing group intervention for women living with HIV: preliminary findings from a randomized controlled trial. Telemed J E Health, 20(2), 128-134. https://doi.org/10.1089/tmj.2013.0072

**17. Martins 2016**

Martins, O. F., Rampal, L., Munn-Sann, L., Sidik, S. M., Othman, N., Iliyasu, Z., & Salawu, F. K. (2016). Effectiveness of Clinician Client Centered Counseling on Condom Use and Status Disclosure of Adult HIV Positive Patients Enrolled In Care in Yola, Nigeria: A Randomized Clinical Trial. International Journal of Health Sciences & Research.

**18. Mashaphu 2020**

Mashaphu, S., Wyatt, G. E., Zhang, M., Mthiyane, T., Liu, H., & Gomo, E. (2020). Effectiveness of an HIV-risk reduction intervention to reduce HIV transmission among serodiscordant couples in Durban, South Africa. A randomized controlled trial. AIDS Care, 32(5), 537-545. https://doi.org/10.1080/09540121.2019.1634785

**19. McKinstry 2017**

McKinstry, L. A., Zerbe, A., Hanscom, B., Farrior, J., Kurth, A. E., Stanton, J., Li, M., Elion, R., Leider, J., Branson, B., & El-Sadr, W. M. (2017). A Randomized-Controlled Trial of Computer-based Prevention Counseling for HIV-Positive Persons (HPTN 065). J AIDS Clin Res, 8(7). https://doi.org/10.4172/2155-6113.1000714

**20. Miller 2019**

Miller, W. C., Rutstein, S. E., Phiri, S., Kamanga, G., Nsona, D., Pasquale, D. K., Rucinski, K. B., Chen, J. S., Golin, C. E., Powers, K. A., Dennis, A. M., Hosseinipour, M. C., Eron, J. J., Chege, W., Hoffman, I. F., & Pettifor, A. E. (2019). Randomized Controlled Pilot Study of Antiretrovirals and a Behavioral Intervention for Persons With Acute HIV Infection: Opportunity for Interrupting Transmission. Open Forum Infect Dis, 6(1), ofy341. https://doi.org/10.1093/ofid/ofy341

**21. Pettifor 2015**

Pettifor, A., Corneli, A., Kamanga, G., McKenna, K., Rosenberg, N. E., Yu, X., Ou, S. S., Massa, C., Wiyo, P., Lynn, D., Tharaldson, J., Golin, C., & Hoffman, I. (2015). HPTN 062: A Pilot Randomized Controlled Trial Exploring the Effect of a Motivational-Interviewing Intervention on Sexual Behavior among Individuals with Acute HIV Infection in Lilongwe, Malawi. PLoS One, 10(5), e0124452. https://doi.org/10.1371/journal.pone.0124452

**22. Samet 2015**

Samet, J. H., Raj, A., Cheng, D. M., Blokhina, E., Bridden, C., Chaisson, C. E., Walley, A. Y., Palfai, T. P., Quinn, E. K., Zvartau, E., Lioznov, D., & Krupitsky, E. (2015). HERMITAGE--a randomized controlled trial to reduce sexually transmitted infections and HIV risk behaviors among HIV-infected Russian drinkers. Addiction, 110(1), 80-90. https://doi.org/10.1111/add.12716

**23. Sarna 2013**

Sarna, A., Luchters, S., Musenge, E., Okal, J., Chersich, M., Tun, W., Mall, S., Kingola, N., & Kalibala, S. (2013). Effectiveness of a community-based positive prevention intervention for people living with HIV who are not receiving antiretroviral treatment: a prospective cohort study. Glob Health Sci Pract, 1(1), 52-67. https://doi.org/10.9745/ghsp-d-12-00023

**24. Sikkema 2014**

Sikkema, K. J., Abler, L., Hansen, N. B., Wilson, P. A., Drabkin, A. S., Kochman, A., MacFarlane, J. C., DeLorenzo, A., Mayer, G., Watt, M. H., & Nazareth, W. (2014). Positive choices: outcomes of a brief risk reduction intervention for newly HIV-diagnosed men who have sex with men. AIDS Behav, 18(9), 1808-1819. https://doi.org/10.1007/s10461-014-0782-3

**25. Smith 2019**

Smith Fawzi, M. C., Siril, H., Liu, Y., McAdam, K., Ainebyona, D., McAdam, E., Somba, M., Oljemark, K., Mleli, N., Lienert, J., Andrew, I., Haberlen, S., Simwinga, A., Todd, J., Makongwa, S., Li, N., & Kaaya, S. (2019). Agents of change among people living with HIV and their social networks: stepped-wedge randomised controlled trial of the NAMWEZA intervention in Dar es Salaam, Tanzania. BMJ Glob Health, 4(3), e000946. https://doi.org/10.1136/bmjgh-2018-000946

**26. Williams 2013**

Williams, J. K., Glover, D. A., Wyatt, G. E., Kisler, K., Liu, H., & Zhang, M. (2013). A sexual risk and stress reduction intervention designed for HIV-positive bisexual African American men with childhood sexual abuse histories. Am J Public Health, 103(8), 1476-1484. https://doi.org/10.2105/ajph.2012.301121

**27. Williams 2012**

Williams, M., Bowen, A., Atkinson, J. S., Nilsson-Schönnesson, L., Diamond, P. M., Ross, M. W., & Pallonen, U. E. (2013). An assessment of brief group interventions to increase condom use by heterosexual crack smokers living with HIV infection. AIDS Care, 24(2), 220-231. https://doi.org/10.1080/09540121.2011.597707

**28. McKirnan 2010**

McKirnan, D. J., Tolou-Shams, M., & Courtenay-Quirk, C. (2010). The Treatment Advocacy Program: a randomized controlled trial of a peer-led safer sex intervention for HIV-infected men who have sex with men. Journal of consulting and clinical psychology, 78(6), 952–963. https://doi-org.libproxy1.nus.edu.sg/10.1037/a0020759

**29. Kalichman 2011**

Kalichman, S. C., Cherry, C., Kalichman, M. O., Amaral, C. M., White, D., Pope, H., Swetzes, C., Eaton, L., Macy, R., & Cain, D. (2011). Integrated behavioral intervention to improve HIV/AIDS treatment adherence and reduce HIV transmission. American journal of public health, 101(3), 531–538. https://doi org. libproxy1. nus.edu.sg/ 10.2105/AJPH.2010.197608

**30. El-Bassel 2010**

El-Bassel, N., Jemmott, J. B., Landis, J. R., Pequegnat, W., Wingood, G. M., Wyatt, G. E., Bellamy, S. L., & NIMH Multisite HIV/STD Prevention Trial for African American Couples Group (2010). National Institute of Mental Health Multisite Eban HIV/STD Prevention Intervention for African American HIV Serodiscordant Couples: a cluster randomized trial. Archives of internal medicine, 170(17), 1594–1601. https://doi-org.libproxy1.nus.edu.sg/10.1001/archinternmed.2010.261

**31. Rose 2010**

Rose, C. D., Courtenay-Quirk, C., Knight, K., Shade, S. B., Vittinghoff, E., Gomez, C., Lum, P. J., Bacon, O., & Colfax, G. (2010). HIV intervention for providers study: a randomized controlled trial of a clinician-delivered HIV risk-reduction intervention for HIV-positive people. Journal of acquired immune deficiency syndromes (1999), 55(5), 572–581. https://doi-org.libproxy1.nus.edu.sg/10.1097/QAI.0b013e3181ee4c62

**32. Olley 2011**

Olley, B., Abbas, M., & Gidron, Y. (2011). The effects of psychological inoculation on cognitive barriers against condom use in women with HIV: A controlled pilot study. SAHARA J : journal of Social Aspects of HIV/AIDS Research Alliance, 8(1), 27–32. https://doi-org.libproxy1.nus.edu.sg/10.1080/17290376.2011.9724981

**33. Rosser 2010**

Rosser, B. R., Hatfield, L. A., Miner, M. H., Ghiselli, M. E., Lee, B. R., Welles, S. L., & Positive Connections Team (2010). Effects of a behavioral intervention to reduce serodiscordant unsafe sex among HIV positive men who have sex with men: the Positive Connections randomized controlled trial study. Journal of behavioral medicine, 33(2), 147–158. https://doi-org.libproxy1.nus.edu.sg/10.1007/s10865-009-9244-1

**34. Chang 2014** *

Chang, L. W., Nakigozi, G., Billioux, V., Serwadda, D., Quinn, T., Bollinger, R., Wawer, M., Gray, R., & Reynolds, S. (2014). Peer support and engagement, HIV care and sexual behaviors among plhiv not on ART: A randomized trial [Conference Abstract]. Topics in antiviral medicine, 22, 561-562. (*This study not included in network but included in the meta-analysis for multiple sexual partners)

#### *Appendix-3: List of references appears to meet the inclusion criteria but not included*

**1. Abdilah 2022**

Abdilah, A., Kadir, H., Mani, K., & Muthiah, G. (2022). Effectiveness of a Safe Sex Education Module in Improving Condom Use among People Living with HIV: A Randomised Controlled Trial [Article]. International Journal of Environmental Research and Public Health, 19(16), Article 10004.

**2. Arends 2022**

Arends, R. M., Grintjes, K. J. T., van den Heuvel, T. J., Foeken-Verwoert, E. G. J., Schene, A. H., van der Ven, A. J. A. M., & Schellekens, A. F. A. (2022). Effectiveness of a group intervention to reduce sexual transmission risk behavior among MSM living with HIV: a non-randomized controlled pilot study [Article]. Aids Care-Psychological and Socio-Medical Aspects of Aids/Hiv, 34(4), 515-526.

**3. Carey 2020**

Carey, M. P., Dunne, E. M., Norris, A. L., Dunsiger, S., Rich, C., Rosen, R. K., Chan, P., & Salmoirago-Blotcher, E. (2020). Telephone-Delivered Mindfulness Training to Promote Medication Adherence and Reduce Sexual Risk Behavior Among Persons Living with HIV: An Exploratory Clinical Trial [Article]. Aids and Behavior, 24(6), 1912-1928.

**4. Chakrapani 2020**

Chakrapani, V., Subramanian, T., Vijin, P. P., Nelson, R., Shunmugam, M., & Kershaw, T. (2020). Reducing sexual risk and promoting acceptance of men who have sex with men living with HIV in India: Outcomes and process evaluation of a pilot randomised multi-level intervention. Glob Public Health, 15(3), 438-451.

**5. Chander 2015**

Chander, G., Hutton, H. E., Lau, B., Xu, X., & McCaul, M. E. (2015). Brief Intervention Decreases Drinking Frequency in HIV-Infected, Heavy Drinking Women: Results of a Randomized Controlled Trial [Article]. Jaids-Journal of Acquired Immune Deficiency Syndromes, 70(2), 137-145.

**6. Chiou 2013**

Chiou, P. Y., Wang, C. C., Chuang, P., Yen, M. Y., & Chang, C. L. (2013). The effect of advanced partner notification for people living with HIV and AIDS [Conference Abstract]. Sexually Transmitted Infections, 89.

**7. Corneli 2014**

Corneli, A., Pettifor, A., Kamanga, G., Golin, C., McKenna, K., Ou, S. S., Hamela, G., Massa, C., Martinson, F., Tharaldson, J., Hilgenberg, D., Yu, X., Chege, W., & Hoffman, I. (2014). HPTN 062: a feasibility and acceptability pilot intervention to reduce HIV transmission risk behaviors among individuals with acute and early HIV infection in Lilongwe, Malawi [Article]. Aids and Behavior, 18(9), 1785-1800.

**8. Hirshfield 2019**

Hirshfield, S., Downing, M. J., Jr., Chiasson, M. A., Yoon, I. S., Houang, S. T., Teran, R. A., Grov, C., Sullivan, P. S., Gordon, R. J., Hoover, D. R., & Parsons, J. T. (2019). Evaluation of Sex Positive! A Video eHealth Intervention for Men Living with HIV [Article]. Aids and Behavior, 23(11), 3103-3118.

**9. Hutton 2015 (abstract)**

Hutton, H. E., Chander, G., Xu, X., & McCaul, M. E. (2015). Brief alcohol intervention reduces unprotected vaginal sex among HIV infected women in care [Conference Abstract]. Alcoholism: Clinical and Experimental Research, 39, 305A.

**10. Jones 2013**

Jones, D. L., Kashy, D., Villar-Loubet, O. M., Cook, R., & Weiss, S. M. (2013). The impact of substance use, sexual trauma, and intimate partner violence on sexual risk intervention outcomes in couples: a randomized trial [Article]. Annals of behavioral medicine: a publication of the Society of Behavioral Medicine, 45(3), 318-328.

**11. Kurth 2016**

Kurth, A. E., Chhun, N., Cleland, C. M., Crespo-Fierro, M., Parés-Avila, J. A., Lizcano, J. A., Norman, R. G., Shedlin, M. G., Johnston, B. E., & Sharp, V. L. (2016). Linguistic and Cultural Adaptation of a Computer-Based Counseling Program (CARE+ Spanish) to Support HIV Treatment Adherence and Risk Reduction for People Living With HIV/AIDS: A Randomized Controlled Trial [Article]. Journal of Medical Internet Research, 18(7), e195.

**12. Lovejoy 2014-1**

Lovejoy, T. I., & Heckman, T. G. (2014). Depression moderates treatment efficacy of an HIV secondary-prevention intervention for HIV-positive late middle-age and older adults. Behav Med, 40(3), 124-133.

**13. Lovejoy 2014-2**

Lovejoy, T. I., Heckman, T. G., & Project, S. I. T. (2014). Telephone-Administered Motivational Interviewing and Behavioral Skills Training to Reduce Risky Sexual Behavior in HIV-Positive Late Middle-Age and Older Adults [Article]. Cognitive and Behavioral Practice, 21(2), 224-236.

**14. Lovejoy 2015**
Lovejoy, T. I., Heckman, T. G., Sikkema, K. J., Hansen, N. B., & Kochman, A. (2015). Changes in Sexual Behavior of HIV-Infected Older Adults Enrolled in a Clinical Trial of Standalone Group Psychotherapies Targeting Depression [Article]. Aids and Behavior, 19(1), 1-8.

**15. Mashaphu 2022**
Mashaphu, S., Wyatt, G. E., Zhang, M., & Liu, H. (2022). Condom use consistency among South African HIV serodiscordant couples following an HIV risk-reduction intervention. Int J STD AIDS, 33(5), 479-484.

**16. Mills 2018**
Mills, E. J., Adhvaryu, A., Jakiela, P., Birungi, J., Okoboi, S., Chimulwa, T. N. W., Wanganisi, J., Achilla, T., Popoff, E., Golchi, S., Karlan, D., Chimulwa, T., & Wangisi, J. (2018). Unconditional cash transfers for clinical and economic outcomes among HIV-affected Ugandan households. AIDS (02699370), 32(14), 2023-2031.

**17. Mirzapour 2022**
Mirzapour, P., Nikoogoftar, M., Dadras, O., SeyedAlinaghi, S., Mohraz, M., & Pirnia, B. (2022). Effectiveness of social marketing on reducing risky sexual behaviors in HIV-positive individuals in Iran: a pilot randomized controlled trial [Article]. HIV and AIDS Review, 21(4), 322-326.

**18. Nostlinger 2016**
Nostlinger, C., Platteau, T., Bogner, J., Buyze, J., Dec-Pietrowska, J., Dias, S., Newbury-Helps, J., Kocsis, A., Mueller, M., Rojas, D., Stanekova, D., van Lankveld, J., Colebunders, R., & Eurosupport Study, G. (2016). Implementation and Operational Research: Computer-Assisted Intervention for Safer Sex in HIV-Positive Men Having Sex With Men: Findings of a European Randomized Multi-Center Trial [Article]. Jaids-Journal of Acquired Immune Deficiency Syndromes, 71(3), E63-E72.

**19. Talebi-Tamijani 2022**
Talebi-Tamijani, Z., Lotfi, R., & Kabir, K. (2022). Tele-counseling based on motivational interviewing to change sexual behavior of women living with HIV: a randomized controlled clinical trial. AIDS Behav, 26(11), 3506-3515.

**20. Lovejoy 2011**
Lovejoy, T. I., Heckman, T. G., Suhr, J. A., Anderson, T., Heckman, B. D., & France, C. R. (2011). Telephone-administered motivational interviewing reduces risky sexual behavior in HIV-positive late middle-age and older adults: a pilot randomized controlled trial. AIDS and behavior, 15(8), 1623–1634.

**21. Tam 2011(abstract)**
Tam, T., Somlak Pedersen, J., Dunham, R., Valle, J., Littlejohn, D., & Tu, D. (2011). Effectiveness of HIV self-management support group program for aboriginal and non-aboriginal peoples living in Vancouver's downtown eastside [Conference Abstract]. Canadian Journal of Infectious Diseases and Medical Microbiology, 22, 16B.

**22. Kalichman 2010**
Kalichman, S. C., Eaton, L., & Cherry, C. (2010). Sexually transmitted infections and infectiousness beliefs among people living with HIV/AIDS: implications for HIV treatment as prevention [Article]. HIV Medicine, 11(8), 502-509.

**23. Rutstein 2015**
Rutstein, S. E., Pettifor, A., Phiri, S., Pasquale, D., Dennis, A., Hosseinipour, M., Kamanga, G., Nsona, D., Hoffman, I., & Miller, W. C. (2015). Pilot study of immediate antiretrovirals and behavioural intervention for persons with acute HIV infection: Opportunity for interrupting transmission [Conference Abstract]. *Sexually Transmitted Infections*, *91*, A77.

**24. Kurth 2016 (abstract)**
Kurth, A., Farrior, J. H., Hanscom, B., McKinstry, L., Stanton, J., Zerbe, A., Elion, R., Leider, J., Branson, B., & El-Sadr, W. M. (2016). Computer-based prevention counseling for HIV-infected persons (HPTN 065) [Conference Abstract]. *Topics in antiviral medicine*, *24*(E-1), 429.

**25. Corneli 2014**
Corneli, A., Pettifor, A., Kamanga, G., Golin, C., McKenna, K., Ou, S. S., Hamela, G., Massa, C., Martinson, F., Tharaldson, J., Hilgenberg, D., Yu, X., Chege, W., & Hoffman, I. (2014). HPTN 062: a feasibility and acceptability pilot intervention to reduce HIV transmission risk behaviors among individuals with acute and early HIV infection in Lilongwe, Malawi [Article]. Aids and Behavior, 18(9), 1785-1800.

#### *Appendix-4: Communication with authors*

61 articles were included in data extraction. However, 22 articles could not be included in the aggregation for various reasons. Therefore, we contacted the authors to request available data,  and the study was excluded if the author did not answer the email in three attempts within 7 days Three of them replied us and two authors provided valid data. In addition, we also found seven pairs of related articles from the same projects, and we retained only the articles with the most informative and complete data.

##### *1) Details of connection*

| Study | Type of data | First connect time | Replied | Provided data | Notes |
|---|---|---|---|---|---|
| Abdilah 2022 | condom usage frequency | 02/09/2023 | no | no | |
| Arends 2022 | Sexual transmission risk behavior (self-defined score) | 02/09/2023 | no | no | |
| Carey 2020 | condom usage frequency | 09/09/2023 | yes | no | No access to data due to retirement |
| Chakrapani 2020 | sexual risk behavior (self-defined score) | 12/09/2023 | no | no | |
| Chander 2015 | changes of unprotected sex (reduced number) | 12/09/2023 | no | no | |
| Chang 2015 | Number of any condom use | 16/09/2023 | no | no | |
| Chiou 2013 (abstract) | condom usage frequency (every time, often, sometimes, never) | NA | no | no | not possible to retrieve a valid email address |

| | | | | | |
|---|---|---|---|---|---|
| Corneli 2013 | Safe sex practices (self-defined score) | 18/09/2023 | no | no | |
| Hirshfield 2019 | changes of unprotected sex (reduced number) | 25/10/2023 | no | no | |
| Hutton 2015(abstract) | Number of unprotected sex | NA | - | - | not possible to retrieve a valid email address |
| Jones 2013 | condom usage frequency | 25/10/2023 | no | no | |
| Kurth 2016 | Sexual transmission risk behavior (self-defined score) | 25/10/2023 | no | no | |
| Lovejoy 2014-1 | Difference in the number of unprotected sex between two groups (OR) | 25/10/2023 | no | no | |
| Lovejoy 2014-2 | number of unprotected sex （all participants, no data for each group） | 25/10/2023 | no | no | |
| Lovejoy 2015 | number of unprotected sex （all participants, no data for each group） | 25/10/2023 | no | no | |

| | | | | | |
|---|---|---|---|---|---|
| Mashaphu 2022 | Percentage of condom use (individual) | 28/10/2023 | no | no | |
| Miller 2019 | The number of condomless sex (no standard deviation) | 19/10/2023 | yes | yes | Author provided the condomless sex acts with the standard deviation for each group |
| Mills 2018 | Condom usage frequency | 19/10/2023 | no | no | |
| Mirzapour 2022 | Sexual risk behavior (self-defined score) | 19/10/2023 | no | no | |
| Martins 2016 | condom use score (self-defined) | 20/10/2023 | yes | yes | Author provided the proportion of condomless sex in each group |
| Nostlinger 2016 | condom use at last intercourse | 22/10/2023 | no | no | |
| Tam 2011 (abstract) | transmission risk behaviors | NA | - | - | not possible to retrieve a valid email address |
| Talebi-Tamijani 2022 | condom use score (self-defined) | 28/10/2023 | no | no | |

### *Appendix-5: Further details on methods*

*1. Data extraction*

1.1 In three-arm or multi-arm studies where the intervention groups differ only in dosage, we included only the results from the group with the higher dosage.

1.2 The "Ycasd" software was used to capture and scale data from graphical representations [3].

1.3 Studies reporting condomless sex within the past 3 months were accepted, while those covering longer period were excluded due to recall bias [4]. We recorded the outcomes as close to 3-month follow up as possible for all analysis [1,2]. If not available, we used data ranging between 2 weeks to 6-month follow up (we gave preference to the timepoint closest to 3-month follow up).

1.4 We extracted both the overall sample size and analytical sample of condomless sex and/or multiple sex partners. The analytical sample was used to compute effective estimates of reducing the condomless sex and multiple sex partners. Discrepancies between the analytical sample and overall sample size were due to missing data.

1.5 In this network meta-analysis, a cluster RCT was included and analyzed alongside individually randomized trials. To account for the inherent intra-cluster correlation and avoid underestimation of variance, we adjusted the effective sample size (ESS) of cluster RCTs using the standard design effect formula:

$$\textbf{Design effect} = 1 + (m - 1) \times \text{ICC}$$

Where m represents the average cluster size and ICC denotes the intraclass correlation coefficient. we assumed an ICC of 0.05 based on prior literature and calculated the effective sample size by applying the design effect. Thus, the number of participants with the outcome and the total number of participants are divided by the design factor and the results analyzed in standard manner with the remaining RCTs.

*2. Data analyses*

2.1 Standard error (SE) and standard deviation (SD)
If a study reported standard error only, we would converse SE to SD using the following formula:

$$SD = SE \times \sqrt{n}$$

2.2 95% confidence interval (CI) and standard deviation (SD)
If a study reported mean and its 95%CI, we would converse 95%CI to SD using the following formula:

$$SD = \frac{Upper\,CI - Lower\,CI}{2 \times Z}$$

Upper CI and Lower CI are the upper and lower limits of the 95% confidence interval.
Z is the Z-score corresponding to the 95% confidence interval, which is approximately 1.96.
If the 95% CI is asymmetric, we would log-transform the mean and 95% CI, and then perform the calculation using the formula.

2.3 Median, minimum, maximum, and first/third quartile of sample
If a study didn't report the mean value but reported the median, minimum, maximum, or first/third quartile of sample, we estimated the mean and standard deviation based on these values using an online calculator (http://www.math.hkbu.edu.hk/~tongt/papers/median2mean.html).

#### *Appendix-6: Characteristics and definition of interventions*

Upon reviewing the categories of intervention, we identified four sexual health interventions namely goal-oriented intervention, education, psychoeducation and safe sex skills training. we identified three types of control conditions in this review, including standard of care, enhanced standard of care [5] and waiting list [6]. Summarily, current interventions emphasize structured approaches, knowledge enhancement, emotional support, and skill development to promote safer sexual behaviors. However, a notable gap remains: none of the existing interventions explicitly incorporate the 'Undetectable = Untransmittable' (U=U) framework. Characteristics and definition of interventions were presented in the Table:

| Type of intervention | Core Elements | Emphasis | Primary Goals | Mechanisms |
|---|---|---|---|---|
| Goal-oriented intervention | • **Assessment**: Identifying specific needs or challenges;<br>• **Goal Setting**: Establishes clear, measurable objectives;<br>• **Planning**: Develops a detailed instructional plan;<br>• **Implementation**: Executes the plan through direct instruction | Structured approach to achieving specific, measurable goals. | Improve safe sexual behaviors by setting and reaching predefined objectives. | Enhances motivation and accountability through a clear, structured plan with specific targets. |
| Education | • **Assessment**: identifying what kind of knowledge is lacking;<br>• **Educational Sessions**: Structured activities providing information on HIV transmission or other STIs prevention | Providing comprehensive knowledge and raising awareness about the consequences of risky sexual behavior. | Enhance understanding and support informed decision-making in relation to sexual behavior. | Increases knowledge and awareness through informative sessions and materials, facilitating better-informed choices. |
| Psychoeducation | • **Assessment**: identifying what kind of knowledge is lacking and needed emotional support;<br>• **Educational and psychological sessions:** offering sexual risk related information and coping strategies. | Integration of psychological and educational techniques to support behavior change | Promote positive cognitive beliefs, healthier practices, and risk understanding. | Combines emotional support with educational content to foster cognitive and behavioral changes. |
| Safe sex skills training | • **Condom Use Practice:** Hands-on training on correct and effective condom use;<br>• **Decision-making and negotiate skills practice**: training on techniques for negotiating condom use with partners and making informed sexual health decisions. | Practical skills for reducing the risk of HIV and STIs during sexual activities | Equip individuals with the knowledge and skills to engage in safer sexual practices. | Provides hands-on practice and skills training to reduce risky behaviors and improve sexual health outcomes |

**Table 1A: Characteristic of 4 sexual health interventions**

| Type of intervention | Number of studies | Definition and Measure |
|---|---|---|
| Goal-oriented intervention (GO) | 13 | GO focuses on setting clear, measurable goals and developing a structured plan to reach them [7,8]. These interventions emphasize the systematic identification of specific needs, the establishment of measurable targets, and the development and execution of a comprehensive strategy that includes instructional methods and resources to support progress towards the goals. |
| Education (Edu) | 8 | Providing structured educational activities includes a variety of components such as educational sessions, psychoeducational materials, HIV transmission information, and more. These activities are designed to enhance knowledge, promote awareness, and support learning. |
| Psychoeducation (PE) | 5 | PE combines psychological and educational techniques, providing coping strategies and supportive counseling. helping individuals promote positive cognitive beliefs, develop healthier practices and understand the risks associated with their behaviors [9,10]. |
| Safe sex skills training (SSST) | 10 | SSST designed to teach individuals the knowledge and skills necessary to engage in sexual activities in a way that reduces the risk of HIV and other sexually transmitted infections (STIs), including condom use practice, condom negotiation skills, decision-making strategies and so on. |
| Standard of care (SOC) | 13 | The term SOC refers to the routine, established practices and treatments that are generally accepted and widely used by the medical community. |
| enhanced Standard of care (eSOC) | 14 | eSOC builds upon the standard of care by incorporating elements such as brief educational sessions, counseling, and other supportive measures. |
| Waiting list (WL) | 5 | Participants who do not receive the experimental treatment, but who are put on a waiting list to receive the intervention after the active treatment group does (from APA dictionary). |

**Table 1B: Definition of included interventions**

#### *Appendix-7: Characteristics of included studies*

**Table 2** Characteristics of included studies (n=34)

| Author year | Area | Participants | Mean age - years (SD) | Sample size (intervention /comparator) | Female (%) | Theoretical basis | Intervention format | Deliverer | Intervention settings | Duration | Follow-up point | Intervention/ comparator |
|---|---|---|---|---|---|---|---|---|---|---|---|---|
| **Bachanas et al 2016 †** | Kenya, Namibia, Tanzania | PLWH | 36.0 | 1778 /1744 | 58.10% | Not stated | Offline; individual | health care providers counselors | HIV clinics | Not stated | 6-month; 12-month | Education / wait list |
| **Brown et al 2019 †** | USA | HIV-infected MSM | 40.6 (7.9) | 39 / 40 | 0% | Information Motivation Behavioral Skills (IMB) Model | Offline; group | MSM facilitators | HIV clinics | 2 240-min sessions, one-off completion | 3-month | Psychoeducation /wait list |
| **Cruess et al 2018 †** | USA | Gay and bisexual men living with HIV | 44· (10.8) | 85 / 82 | 0% | Information Motivation Behavioral Skills (IMB) mode | Online; group | Counselors | Free online open-source survey service | 2 45-min sessions, once a week for 2 weeks | 6-month | Education/ enhanced standard of care |
| **Dévieux et al 2022 †** | USA | Alcohol-using PLWH | 44.8 (7.2) | 160 / 161 | 34.20% | Information Motivation Behavioral Skills (IMB) mode | Offline; group | Counselors | Community-Based Organizations | 8 sessions; twice a week for 4 weeks | 3-month; 6-month; 12-month | Psychoeducation /standard of care |
| **Echenique et al 2013 †** | USA | Women living with HIV | Not stated | 65 / 41 | 100% | Not stated | Offline; individual | Not stated | HIV clinics | Four sessions, once a week for 4 weeks | 6-month; 12-month | Safe sex skills training / standard of care |
| **Smith et al 2019 *†** | Tanzania | PLWH | Not stated | 224 / 548 | 57.00% | Not stated | Offline; group | Health workers | HIV care and treatment center (CTC) | 10 sessions, once a week for 10 weeks | 6-month | Psychoeducation /standard of care |
| **Fisher et al 2014 †** | South Africa | PLWH | 37.3 (9.0) | 924 / 967 | 55.60% | Information Motivation Behavioral Skills (IMB) model | Offline; individual | Lay counselor | HIV clinical care sites | 10-15 min counseling integrated into PLWH's routine clinical care; | 6-month; 12-month; 18-month | Goal-oriented intervention/standard of care |
| **McKirnan et al 2010†** | USA | MSM living with HIV | 42 | 165 /148 | 0% | Not stated | Online; individual | Counselor | HIV Clinic | 4 60-90 min counselings within 3 months | 6-month; 12-month | Goal-oriented intervention/waiting list |
| **Kalichman et al 2011†** | USA | PLWH | 44.1 (6.8) | 217 /219 | 45% | Conflict theory (decision making) | Offline; group and individual | Facilitators | AIDS service Center | 5 120 min group sessions and a 60 min one to one counseling | 3-month; 6-month; 9-month | Goal-oriented intervention/ enhanced standard of care |
| **Golin et al 2013 †** | USA | PLWH | 42.7 (9.1) | 242 / 248 | 25% | Social cognitive theory | Offline and online; individual | Counselor | HIV clinics | 4 (MI session+5 CD sessions) once a month for 4 months | 4-month; 8-month; 12-month | Goal-oriented intervention/ enhanced standard of care |
| **Hart et al 2021 †** | Canada | Gay and Bisexual Men Living With HIV | 40.8 (10.9) | 89 / 93 | 0% | Not stated | Offline; group | Peers | Community-Based Organizations | 8 120 min sessions; once a week for 2 months | 3-month; 6-month | Goal-oriented intervention/standard of care |

**Table 2** (Continued)

| Author year | Area | Participants | Mean age - years (SD) | Sample size (intervention / comparator) | Female (%) | Theoretical basis | Intervention format | Deliverer | Intervention settings | Duration | Follow-up point | Intervention/ comparator |
|---|---|---|---|---|---|---|---|---|---|---|---|---|
| Holstad et al 2012 *† | Nigeria | HIV positive women | 30.7 (5.9) | 28 / 20 | 100% | Social cognitive theory | Offline; group | Facilitators | HIV Clinic | 8 90-120 min sessions; once a week for 2 months | 6-month | Safe sex skills training / enhanced standard of care |
| Kahler et al 2018 † | USA | Alcohol use in HIV-infected MSM | 42.1 (10.4) | 79 / 88 | 0% | Not stated | Offline and online; individual | Counselor | HIV Clinic | Single 60 min counseling and two 20 min counseling | 3-month; 6-month; 12-month | Goal-oriented intervention /standard of care |
| Kalichman et al 2018 | USA | PLWH | 44.5 (10.2) | 229 / 236 | 23.40% | Information Motivation Behavioral Skills (IMB) model | Offline and online; group and individual | Facilitators | HIV Clinic and cell phone | 4 cellphone sessions(bi-weekly) and a 120 min group session | at 2,4,5,6,7,9,10,1 1,12-month | Safe sex skills training / enhanced standard of care |
| Klein et al 2013 *† | USA | African-American women | 40.7 (8.6) | 81 / 87 | 100% | Social cognitive theory | Online; individual | Self help | mHealth | 27 2-8min activity modules; self-paced learning within a month | 3-month | Safe sex skills training / enhanced standard of care |
| McKinstry et al 2017 † | USA | PLWH | 45.0 (10.3) | 441 / 453 | 30% | Information Motivation Behavioral Skills (IMB) model; the Transtheoretical model of change; Social cognitive behavioral theory | Online; individual | Self help | mHealth | 4 sessions; once every 3 months for 1 year | 3-month; 6-month; 12-month | Goal-oriented intervention / standard of care |
| Kurth et al 2014 † | USA | PLWH | 51.4(a) | 111 / 113 | 8.79% | Information Motivation Behavioral Skills (IMB) model; the Transtheoretical model of change; Social cognitive behavioral theory | Online; individual | Self help | mHealth | 12 sessions; four times every 3 months for 9 months | 3-month; 6-month; 9-month; 12-month | Goal-oriented intervention/standard of care |
| Liu et al 2018 † | China | Newly-infected HIV positive MSM | 28.0(a) | 184 / 183 | 0% | Information Motivation Behavioral Skills (IMB) model | Offline; individual | Peers | Community-based organization | 4 60 min counseling; scheduled visit within 7 days | 3-month; 6-month; 9-month; 12-month | Goal-oriented intervention/standard of care |
| Marhefka et al 2014 † | USA | Women Living with HIV | 42.8 (8.2) | 36 / 35 | 100% | ADAPT-ITT model | Offline; group | Facilitators | Community-based organization | 6 120 min sessions; twice a week for 3 weeks | 6-month | Safe sex skills training /wait list |
| Mashaphu et al 2020 † | Durban | HIV-infected serodiscordant couples | 39.3 (9.2) | 20 / 10 | NA | Not stated | Offline; group | Psychiatrist | HIV Clinic | 3 120min sessions; once every 4 weeks for 3 months | 1-month | Safe sex skills training /wait list |

**Table 2** (Continued)

| Author year | Area | Participants | Mean age - years (SD) | Sample size (intervention / comparator) | Female (%) | Theoretical basis | Intervention format | Deliverer | Intervention settings | Duration | Follow-up point | Intervention/ comparator |
|---|---|---|---|---|---|---|---|---|---|---|---|---|
| Milam et al 2016 † | USA | HIV positive MSM | 43.7 | 90 / 89 | 0% | Social cognitive behavioral theory; the Transtheoretical model of change | Online; individual | Self help | mHealth | 4 sessions every month for 12 months | at 1, 2,4,5,6,7,8,9,10, 11,12-month | Education/enhanced standard of care |
| Miller et al 2019 † | Malawi | Newly HIV infection individuals | 28.0 | 18 / 9 | 39% | Not stated | Offline; group | Counselor | HIV Clinic | 5 sessions within 2months | 26-week; 52-week | Goal-oriented intervention/standard of care |
| Martins et al 2016 † | Nigeria | PLWH | 37.2 (9.8) | 114 / 123 | 72.50% | Information Motivation Behavioral Skills (IMB) model | Offline; individual | Counselor | HIV Clinic | 2 10-15 min counseling; once a month for 2 months | 2-month; 6-month | Goal-oriented intervention/standard of care |
| Pettifor et al 2015 † | Malawi | Newly HIV infection individuals | Not stated | 14 / 13 | 33.30% | Information Motivation Behavioral Skills (IMB) model | Offline; individual | Counselor | HIV Clinic | 4 counseling within 2 weeks and a booster at 8th week | 24-week | Education/enhanced standard of care |
| Samet et al 2015 † | Russia | HIV-infected Russian Drinkers | 30.1 (5.1) | 264 / 259 | 40.70% | Social Cognitive Theory | Offline; individual and group | Psychologists and physicians | HIV Clinic | 5 sessions including 2 individual sessions and 3 group sessions | 6-month | Education/enhanced standard of care |
| Sarna et al 2013 *† | Kenya | PLWH | 35.4 (8.3) | 325 / 309 | 74.30% | Not stated | Offline; individual | Community health workers | Community-based organization | 4 30-60 min sessions within 6 months | 6-month; 12-month | Goal-oriented intervention/standard of care |
| Sikkema et al 2014 † | USA | Newly-infected HIV positive MSM | 32.3 (8.2) | 38 / 41 | 0% | Information Motivation Behavioral Skills (IMB) model; the Transtheoretical model of change | Offline; individual | Counselor | Federally Qualified Health Center | 3 60 min sessions within a month | 3-month; 6-month; 9-month; | Goal-oriented intervention/standard of care |
| Williams et al 2013 *† | USA | HIV-Positive Bisexual Men | 46.6 (8.3) | 44 / 44 | 0% | Cognitive Behavioral Theory | Offline; group | Facilitator | HIV Clinic | 6 120 min sessions; twice a week for 3 weeks | 3-month; 6-month | Psychoeducation /enhanced standard of care |
| Williams et al 2012 † | USA | HIV-Positive heterosexual Men | 43.3 | 165 / 182 | 53% | Social cognitive theory; the integrative model of behavioral prediction | Offline; group | Community health workers | Community-based organization | 6 60 min sessions within 4 weeks | 3-month; 9-month | Education/enhanced standard of care |
| Chang et al 2015 *† | Rakai, Uganda | PLWH | 30.0 | 170 / 174 | 63% | Information Motivation Behavioral Skills (IMB) model | Offline; group | Peers | Home-based | 12 monthly home-based counseling | 12-month | Psychoeducation /enhanced standard of care |

**Table 2** (Continued)

| Author year | Area | Participants | Mean age - years (SD) | Sample size (intervention / comparator) | Female (%) | Theoretical basis | Intervention format | Deliverer | Intervention settings | Duration | Follow-up point | Intervention/ comparator |
|---|---|---|---|---|---|---|---|---|---|---|---|---|
| **El-Bassel et al 2010†** | USA | HIV Serodiscordant Couples | 43 | 232 / 235 | NA | Social cognitive theory | Offline; group and individual | Cofacilitators | HIV Clinic | 8 120 min sessions once a week | postintervention ; 6-month; 12-month | Safe sex skills training / enhanced standard of care |
| **Kalichman et al 2011†** | USA | PLWH | 43 | 181 / 205 | 26.7% | theory of planned behavior and the information-motivation-behavioral skills (IMB) model | Offline; group | Community health workers (CHWs) | HIV Clinic | 2 120-min group sessions and a booster session at 4 weeks. | 6-month | Safe sex skills training / standard of care |
| **Benjamin et al 2011†** | Africa | Women Living with HIV | 28.6 (4.2) | 10 / 11 | 100% | Not stated | Offline; group | Facilitator | HIV Clinic | 4 20-min sessions | postintervention | psychoeducation / education |
| **Rosser et al 2010†** | USA | MSM Living with HIV | Not stated | 166 / 194 | 0% | Not stated | offline; group | Community health workers (CHWs) | Community-based organization | one-weekend, 14- to 16-hour structured sessions | 6-month; 12-month; 18-month | Safe sex skills training /education |

**Note:** RCT, Randomized controlled trial; PLWH, People living with HIV; MSM, men who have sex with men; SD, standard deviation; Age labeled with (a) indicates the median. * studies reported multiple sexual partners; † studies reported condomless sex.

***Appendix-8: Summary of risk of bias***

| | Randomization process | Deviations from the intended interventions | Missing outcome data | Measurement of the outcome | Selection of the reported result | Overall |
|---|---|---|---|---|---|---|
| Bachanas 2016 | ! | + | ! | + | + | ! |
| Brown 2019 | ! | ! | ! | + | ! | ! |
| Cruess 2018 | + | + | ! | + | + | ! |
| Dévieux 2022 | + | ! | ! | + | ! | ! |
| Echenique 2013 | ! | ! | ! | + | + | ! |
| Smith 2019 | + | + | + | + | + | + |
| Fisher 2014 | + | - | ! | + | + | - |

+ Low risk

! Some concerns

- High risk

| | | | | | | |
|---|---|---|---|---|---|---|
| Golin 2012 | ! | ! | ! | + | + | ! |
| Hart 2021 | ! | ! | + | + | ! | ! |
| Holstad 2012 | + | ! | - | ! | + | - |
| Kahler 2018 | + | ! | ! | ! | + | ! |
| Kalichman 2018 | + | + | ! | ! | ! | ! |
| Klein 2013 | ! | + | - | - | ! | - |
| McKinstry 2017 | + | + | ! | ! | + | ! |
| Kurth 2014 | ! | + | - | - | ! | - |
| Liu 2018 | + | + | + | + | + | + |
| Marhefka 2014 | + | + | + | ! | + | ! |
| Mashaphu 2020 | ! | ! | ! | + | + | ! |
| Milam 2016 | ! | ! | - | + | ! | - |
| Miller 2019 | ! | ! | ! | + | ! | ! |
| Martins 2016 | ! | + | ! | ! | ! | ! |

| | | | | | | |
|---|---|---|---|---|---|---|
| Pettifor 2015 | + | - | - | ! | - | - |
| Sarna 2013 | ! | ! | - | ! | - | - |
| Samet 2015 | - | - | ! | + | + | - |
| Sikkema 2014 | ! | ! | ! | + | + | ! |
| Williams 2013 | + | + | + | + | + | + |
| Williams 2012 | + | ! | - | - | ! | - |
| McKirnan 2010 | + | - | + | + | ! | ! |
| Kalichman 2011 | + | ! | + | + | + | ! |
| El-Bassel 2010 | + | ! | + | + | + | ! |
| Rose 2010 | - | ! | - | + | + | - |
| Benjamin 2011 | ! | ! | + | + | - | - |
| Rosser 2010 | + | + | + | + | + | + |
| Chang 2015 | + | ! | ! | ! | ! | ! |

Note: rules of ROB 2.0 for assessing risk of bias
(A=low risk, B=high risk, C=some concerns)

**1. Randomization process**

A: The researcher describes the random component of the sequence generation process, such as using a random number table, flipping a coin, shuffling cards, or

sealing envelopes.
B: The researcher describes non-random components of the sequence generation process, such as parity based on date of birth, algorithms based on date, hospital or clinic record numbers, etc.
C: There is insufficient information to determine the sequence generation process.
**2. Deviations from the intended interventions**
A: Participants and researchers were unable to predict allocation (e.g., using centralized allocation; or using consecutively numbered, opaque, and sealed envelopes).
B: Participants and researchers were able to predict the impending allocation (e.g., open random allocation table, list of random numbers); or the envelopes were unsealed, opaque, or not sequentially numbered.
C: Insufficient information to determine the circumstances under which the allocation was hidden or the method was not described.
**3. Measurement of the outcome**
A: Blinding of participants, key investigators, and outcome assessors was implemented and the blinding was unlikely to be broken. Alternatively, blinding is not implemented but is unlikely to introduce bias. Unblinded and unlikely to introduce bias.
B: Blinding was not implemented or was incomplete and the outcome may have been influenced by the lack of blinding.
C: Insufficient information to judge the adequacy of blinding.
**4. Missing outcome data**
A: No missing outcome data, the reason for missing data may not be related to the true outcome, or the amount of missing data is balanced between groups.
B: Reasons for missing outcome data may be related to the true outcome and the amount of data is not balanced between groups or there are differences in the reasons for missing data.
C: Data on missing visits or exclusions are underreported.
**5. Selection of the reported result**
A: The trial protocol provided clearly aligns the primary outcome with the final trial report.
B: Primary outcome is not consistent between the trial protocol and the final trial report.
C: No trial protocol available or insufficient reporting to determine whether selective reporting exists.

#### *Appendix-9: Assessment of Model fit*

Totresdev: posterior mean total residual deviance.

DIC:  deviance information criterion.

Convergence: number of iterations before convergence occurred.

| Outcomes | Model | Datapoints | Totresdev | DIC | Convergence |
|---|---|---|---|---|---|
| Proportion-proportion of condomless sex | Random effects | 42 | 44.15 | 77.68 | 150,000 |
| | Fixed effects | | 54.79 | 82.18 | 150,000 |
| Frequency-mean number of condomless sex in the past month | Random effects | 28 | 28.41 | 54.96 | 150,000 |
| | Fixed effects | | 63.71 | 83.79 | 150,000 |

***Appendix-10: Summary Estimates from Pair-wise Meta-analysis of Direct Comparisons***

*1) Proportion- proportion of condomless sex*

| Format Comparison | No. | OR (95% CI) | $I^2$ statistics | Egger Test *P* value |
|---|---|---|---|---|
| SSST vs | | | | |
| eSOC | 2 | 0.76 (0.57, 1.02) | 0.00% | NC |
| SOC | 2 | 0.91 (0.59, 1.40) | 0.00% | NC |
| WL | 2 | 0.85 (0.43, 1.67) | 0.00% | NC |
| Edu | 1 | 0.94 (0.61, 1.47) | NC | NC |
| Edu vs | | | | |
| SOC | 1 | **0.63 (0.50, 0.80)** | NC | NC |
| eSOC | 4 | 1.00 (0.80, 1.25) | 7.10% | 0.400 |
| GO vs | | | | |
| SOC | 6 | **0.50 (0.34, 0.74)** | 64.9% | 0.972 |
| WL | 1 | 0.86 (0.55, 1.36) | NC | NC |
| PE vs | | | | |
| SOC | 1 | 0.99 (0.72, 1.37) | NC | NC |
| Edu | 1 | 0.73 (0.21, 2.57) | NC | NC |

Notes: WL = waiting list; SOC = standard of care; eSOC = enhanced standard of care; PE = psychoeducation; SSST = safe sex skills training; Edu = education.

*2) Frequency- mean number of condomless sex in the past month*

| Format Comparison | No. | SMD (95% CI) | $I^2$ statistics | Egger Test *P* value |
|---|---|---|---|---|
| PE vs | | | | |
| eSOC | 1 | 0.02 (-0.39, 0.44) | NC | NC |
| SOC | 1 | -0.17 (-0.39, 0.04) | NC | NC |
| WL | 1 | -0.30 ( -0.75, 0.13) | NC | NC |
| SSST vs | | | | |
| WL | 1 | **-1.05 (-1.55, -0.55)** | NC | NC |
| eSOC | 2 | -0.05 (-0.57, 0.46) | 93.8% | NC |
| GO vs | | | | |
| SOC | 4 | **-0.12 (-0.20, -0.03)** | 0.00% | 0.222 |
| eSOC | 3 | -0.25 (-0.51, 0.02) | 77.7% | 0.071 |
| Edu vs | | | | |
| eSOC | 1 | -0.12 (-0.42, 0.18) | NC | NC |

Notes: GO = goal-oriented intervention; WL = waiting list;
SOC = standard of care; eSOC = enhanced standard of care; PE = psychoeducation; SSST = safe sex skills training; Edu = education.

#### *Appendix-11: Network rank test*

##### *1. Results of SUCRA and mean rank*

| Treatment | Proportion (mean rank) | Frequency (mean rank) |
|---|---|---|
| GO | 0.903(1.6) | 0.828 (2.0) |
| Edu | 0.584 (3.5) | 0.651 (3.1) |
| PE | 0.422 (4.5) | 0.435 (4.4) |
| SSST | 0.727 (2.6) | 0.707 (2.8) |
| eSOC | 0.167 (6.0) | 0.472 (4.2) |
| WL | 0.518 (3.9) | 0.015(6.9) |
| SOC | 0.179 (5.9) | 0.392 (4.6) |

Notes: SUCRA = the surface under the cumulative ranking curves; GO = goal-oriented intervention; WL = waiting list; SOC = standard of care; eSOC = enhanced standard of care; PE = psychoeducation; SSST = safe sex skills training; Edu = education.

##### *2. Cumulative probability plots*

##### *1) Proportion*

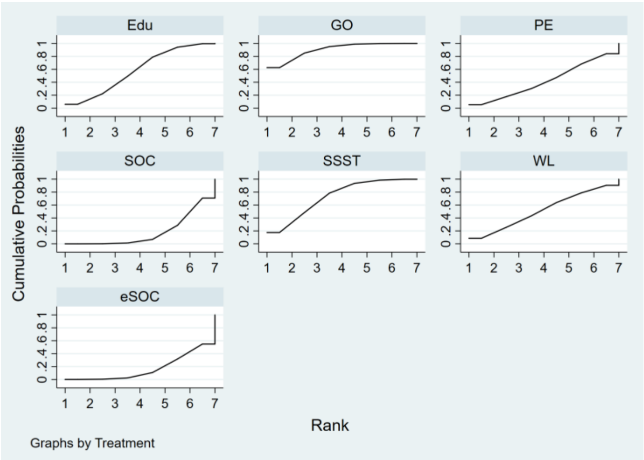

##### *2)Frequency*

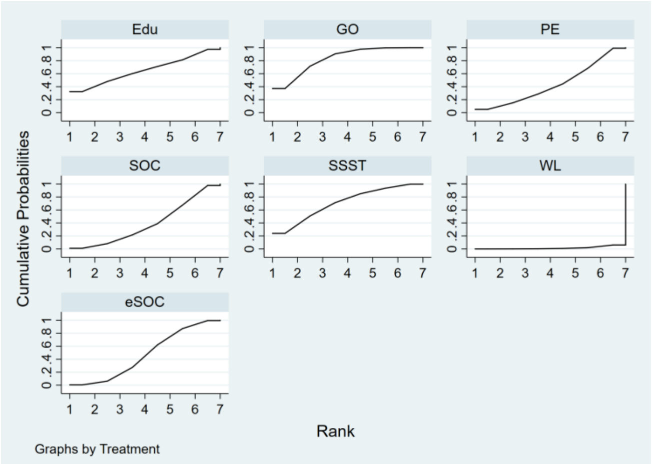

#### *Appendix-12: Subgroup analyses for GO and SSST*

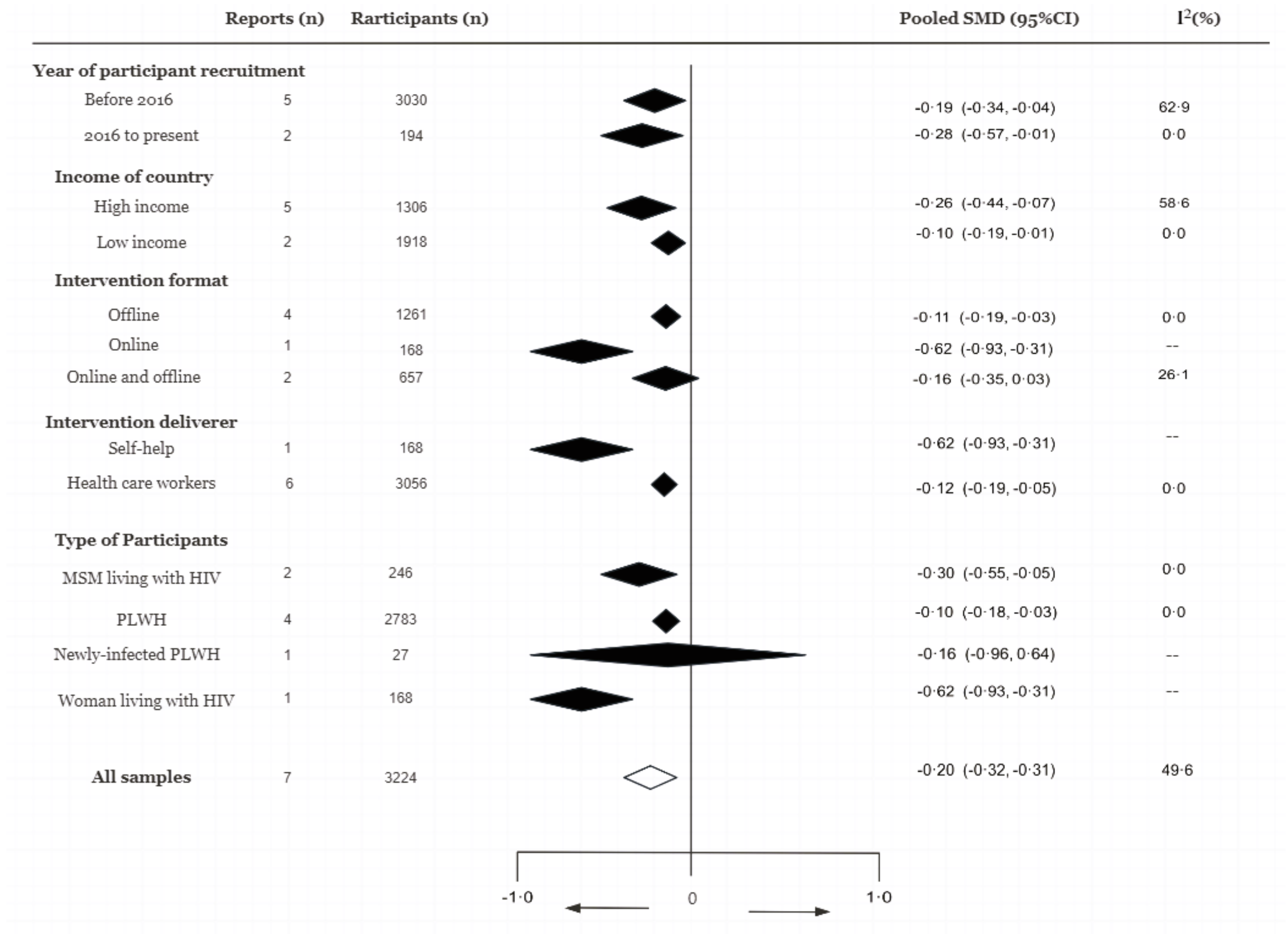

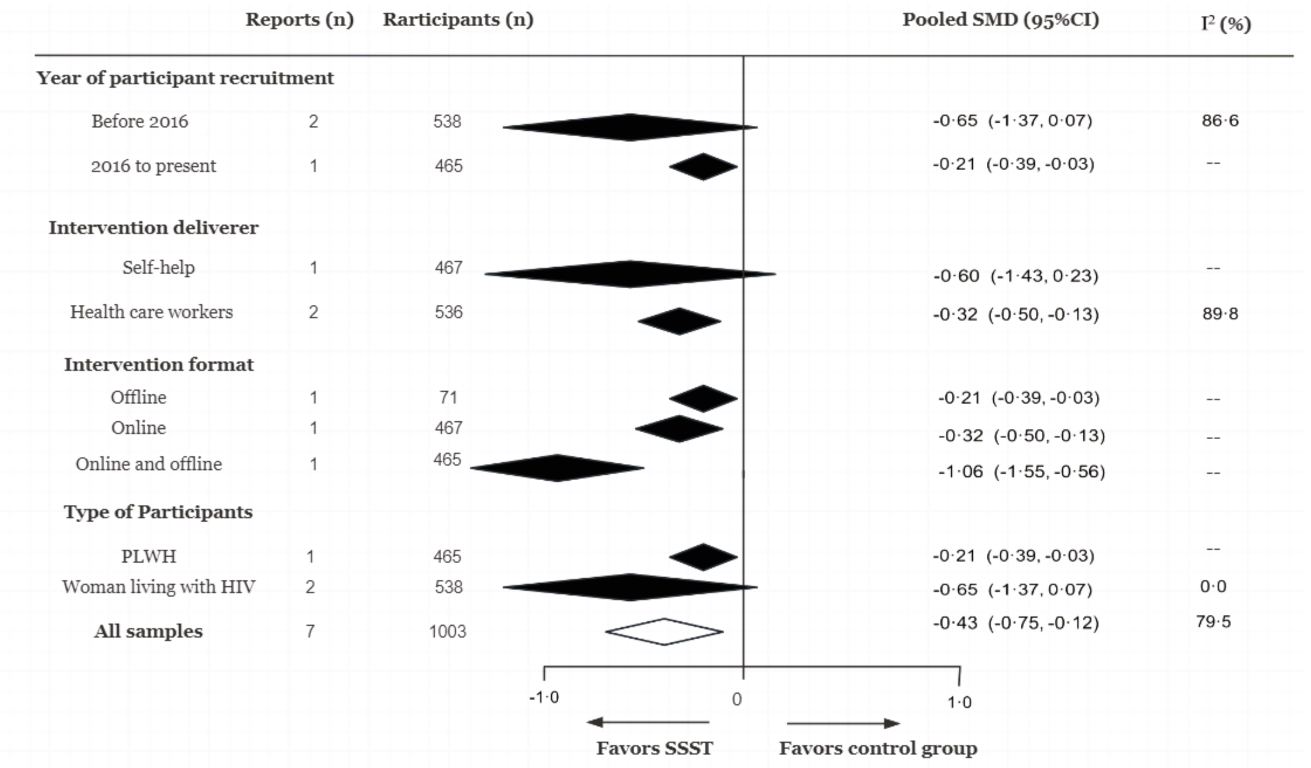

**Figure. subgroup meta-analysis of the effectiveness of goal-oriented intervention and safe sex skills training for reducing the mean number of condomless sex in the past month**

SMD= standard mean differences; PLWH=people living with HIV; MSM= men who have sex with men. GO=goal-oriented intervention; SSST=safe sex skills training.

*Appendix-13: Assessment of Transitivity*

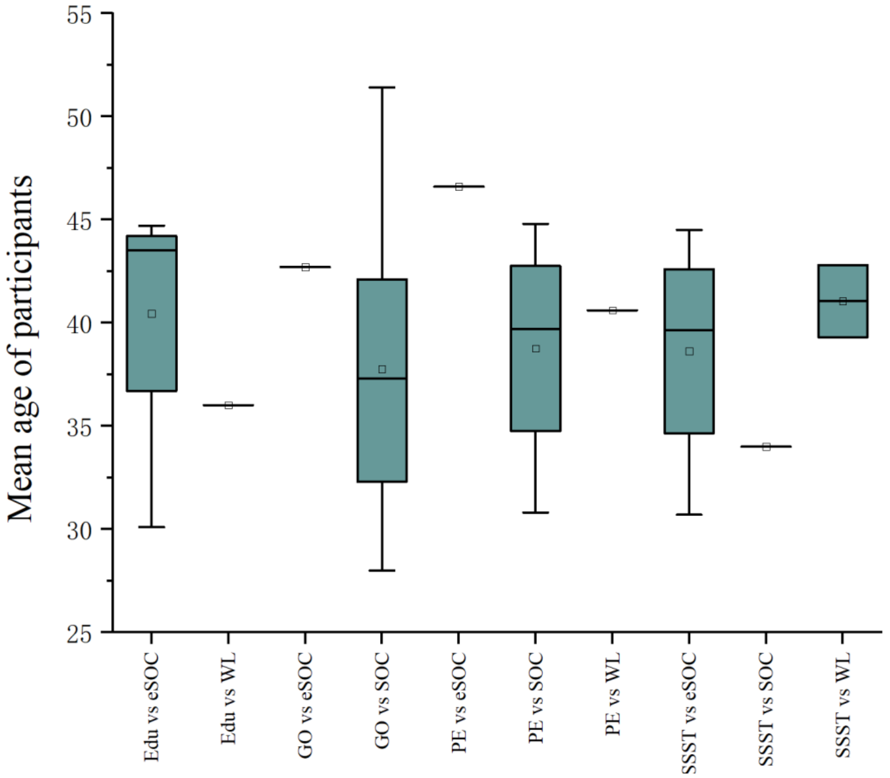

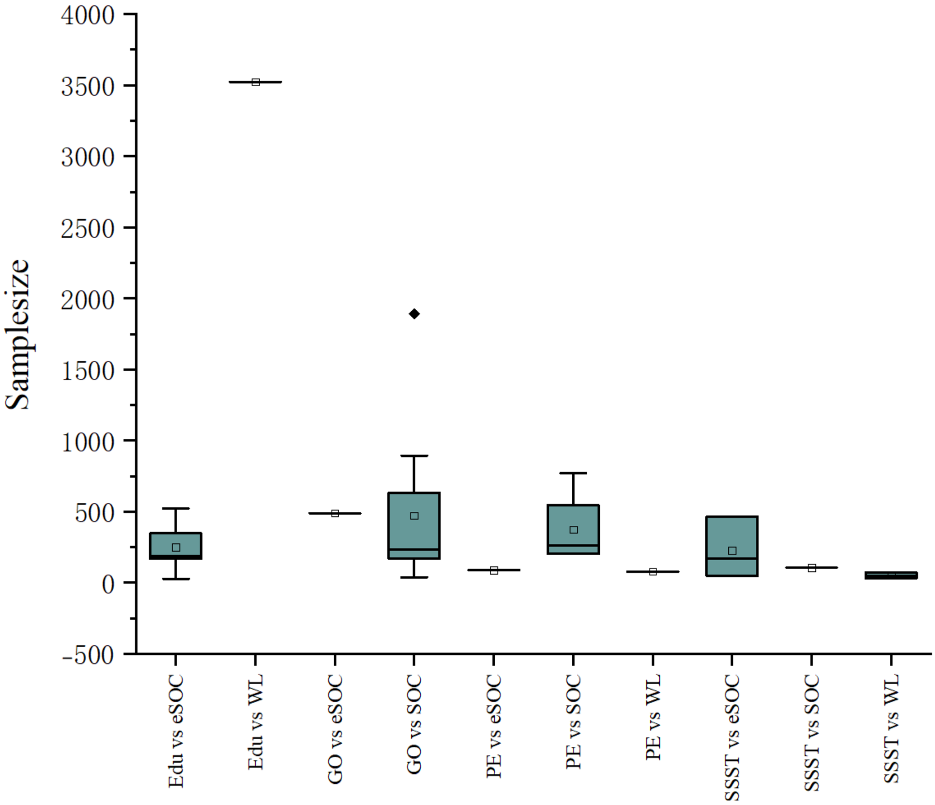

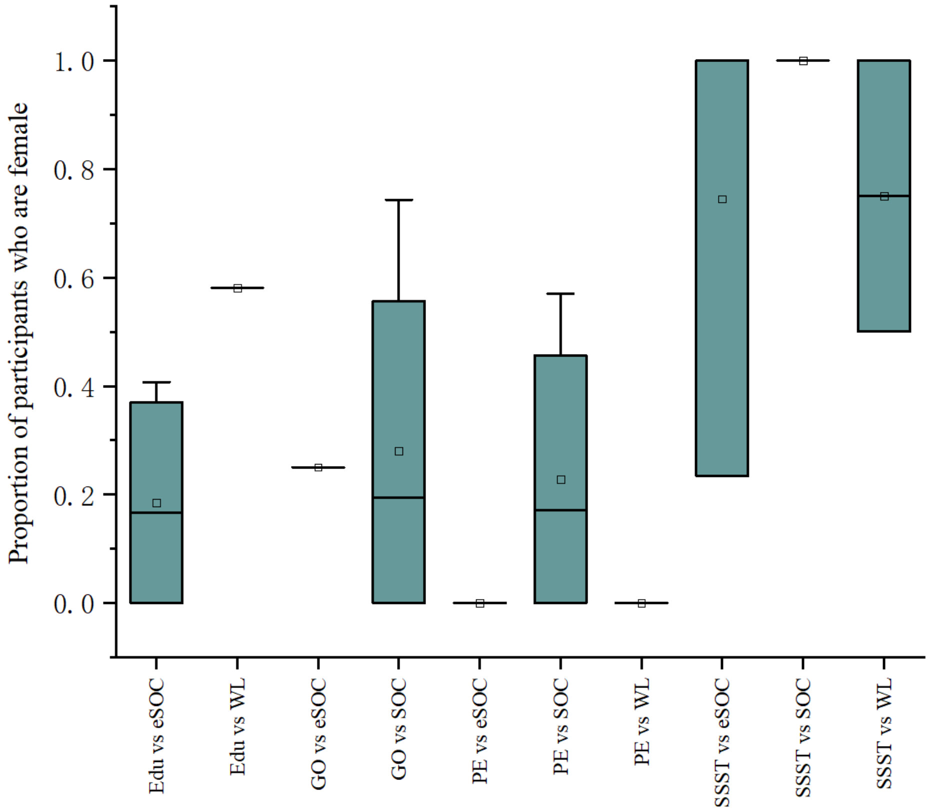
Proportion of participants who are female
0.0
0.2
0.4
0.6
0.8
1.0
Edu vs eSOC
Edu vs WL
GO vs eSOC
GO vs SOC
PE vs eSOC
PE vs SOC
PE vs WL
SSST vs eSOC
SSST vs SOC
SSST vs WL

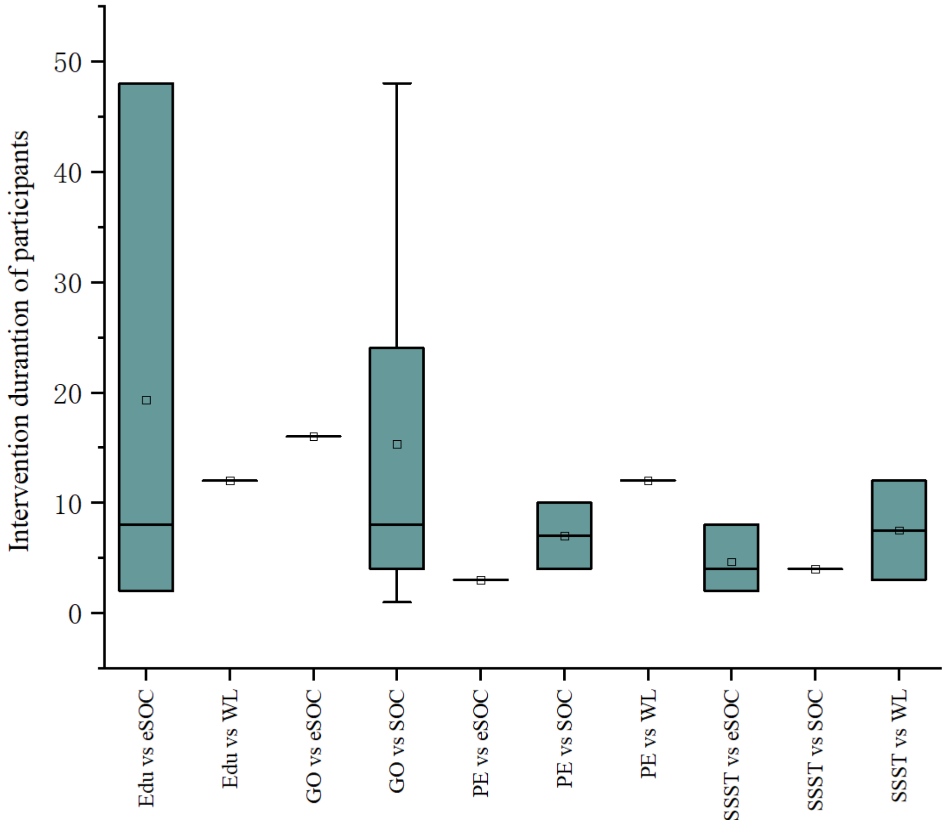
Intervention durantion of participants
0
10
20
30
40
50
Edu vs eSOC
Edu vs WL
GO vs eSOC
GO vs SOC
PE vs eSOC
PE vs SOC
PE vs WL
SSST vs eSOC
SSST vs SOC
SSST vs WL

#### *Appendix-14: Assessment of Heterogeneity*

We assessed the extent of heterogeneity by comparing the estimated tau2 for between-study variability with the empirical distributions for tau as provided by Turner et al. for proportion of condomless sex and Rhodes et al. for number of condomless sex (in the past month).

We judged heterogeneity as very low when below the 25% quantile, as low when between 25% and 50% quantile, as moderate when between 50% and 75% and as high when above the 75% quantile.

| Outcomes | Between study variance (tau) | Outcome type used as comparator | Predictive distribution of tau -- Median (IQR) | Judgement on heterogeneity |
|---|---|---|---|---|
| Proportion of condomless sex | 0.23 | Subjective outcomes (various) —from [11] | 0.38 (0.36, 0.42) | Very Low |
| Mean number of condomless sex (in the past month) | 0.19 | Various subjectively measured outcomes —from [12] | 0.26 (0.026, 3.00) | Low |

***Appendix-15: Assessment of Inconsistency***

*1. Proportion- proportion of condomless sex*

*1.1 Loop-specific approach*

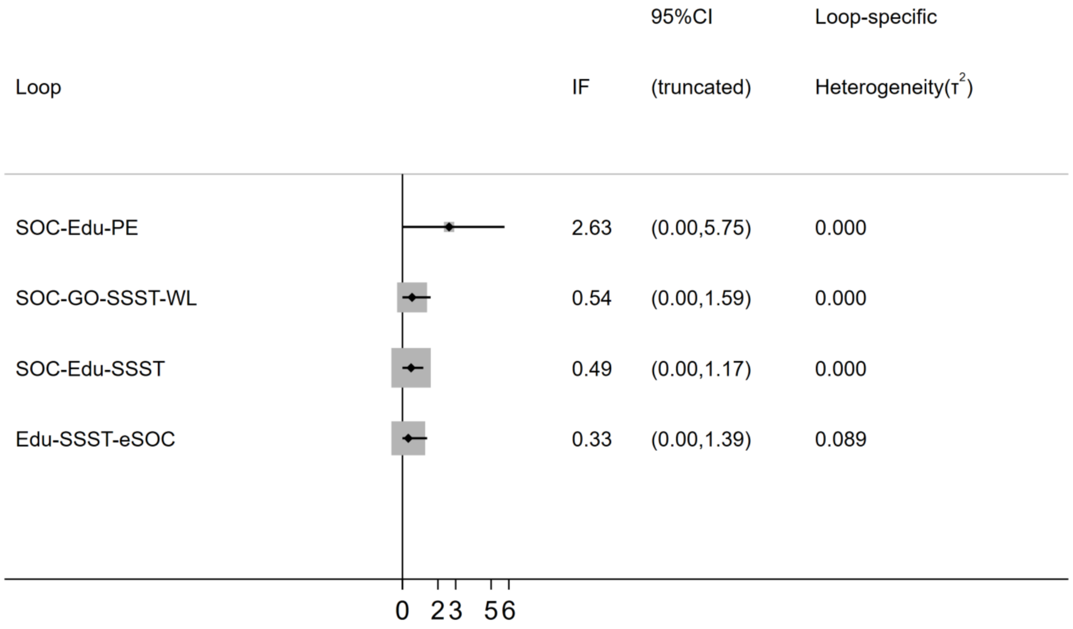

| Loop | IF | seIF | z_value | p_value | CI_95 | Loop_Heterog_tau2 |
|---|---|---|---|---|---|---|
| SOC-Edu-PE | 2.63 | 1.59 | 1.65 | 0.09 | (0.00,5.75) | 0.000 |
| SoC-Edu-SSST | 0.48 | 0.35 | 1.38 | 0.16 | (0.00,1.17) | 0.000 |
| Edu-SSST-eSOC | 0.33 | 0.54 | 0.61 | 0.54 | (0.00,1.39) | 0.000 |
| SOC-GO-SSST-WL | 0.54 | 0.53 | 1.01 | 0.31 | (0.00,1.59) | 0.089 |

*1.2 Side-splitting approach*

| Side | Direct | | Indirect | | Difference | | P>z | tau |
|---|---|---|---|---|---|---|---|---|
| | Coef. | Std. Err. | Coef. | Std. Err. | Coef. | Std. Err. | | |
| SOC GO | -0.64 | 0.18 | -0.33 | 0.61 | -0.31 | 0.64 | 0.63 | 0.25 |
| SOC Edu | -0.53 | 0.24 | 0.09 | 0.33 | -0.62 | 0.41 | 0.13 | 0.21 |
| SOC PE | -0.11 | 0.25 | -2.51 | 1.61 | 2.40 | 1.63 | 0.14 | 0.23 |
| SOC SSST | -0.14 | 0.29 | -0.72 | 0.28 | 0.58 | 0.40 | 0.15 | 0.20 |
| WL GO | 0.24 | 0.36 | 0.55 | 0.53 | -0.31 | 0.64 | 0.63 | 0.25 |
| Edu PE | -2.16 | 1.60 | 0.24 | 0.33 | -2.40 | 1.63 | 0.14 | 0.23 |
| Edu SSST | -0.08 | 0.35 | -0.15 | 0.27 | 0.07 | 0.44 | 0.88 | 0.26 |
| eSOC Edu | 0.23 | 0.18 | 0.71 | 0.35 | -0.49 | 0.40 | 0.23 | 0.21 |
| eSOC SSST | 0.66 | 0.26 | 0.17 | 0.30 | 0.49 | 0.40 | 0.23 | 0.21 |
| WL SSST | 0.29 | 0.43 | -0.02 | 0.47 | 0.31 | 0.64 | 0.63 | 0.25 |

Notes: GO = goal-oriented intervention; WL = waiting list; SOC = standard of care; eSOC = enhanced standard of care; PE = psychoeducation; SSST = safe sex skills training; Edu = education.

*1.3 Design-by-treatment test*

chi2 (10) = 1.27
Prob > chi2 = 0.3395

#### *2. Frequency- mean number of condomless sex in the past month*

##### *2.1 Loop-specific approach*

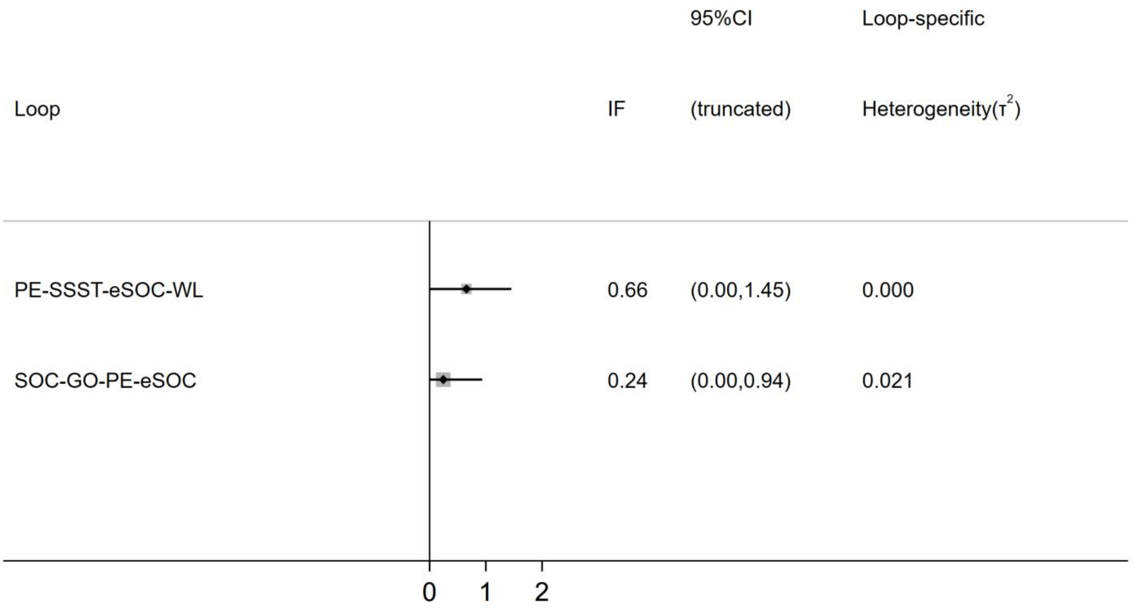

| Loop | IF | seIF | z_value | p_value | CI_95 | Loop_Heterog_tau2 |
|---|---|---|---|---|---|---|
| PE-SSST-eSOC-WL | 0.65 | 0.41 | 1.61 | 0.11 | (0.00,1.45) | 0.000 |
| SOC-GO-PE-eSOC | 0.31 | 0.41 | 0.75 | 0.49 | (0.00,0.94) | 0.021 |

##### *2.2 Side-splitting approach*

| Side | Direct | | Indirect | | Difference | | P>z | tau |
|---|---|---|---|---|---|---|---|---|
| | Coef. | Std. Err. | Coef. | Std. Err. | Coef. | Std. Err. | | |
| SOC GO | -0.21 | 0.15 | -0.64 | 0.40 | 0.43 | 0.42 | 0.31 | 0.23 |
| SOC PE | -0.17 | 0.26 | 0.26 | 0.34 | -0.43 | 0.42 | 0.31 | 0.23 |
| eSOC GO | 0.25 | 0.15 | -0.18 | 0.40 | 0.43 | 0.42 | 0.31 | 0.23 |
| eSOC Edu | 0.12 | 0.28 | -0.24 | 199.89 | 0.36 | 199.89 | 1.00 | 0.23 |
| eSOC PE | -0.03 | 0.33 | -0.06 | 0.29 | 0.03 | 0.44 | 0.94 | 0.25 |
| WL PE | 0.30 | 0.31 | 1.08 | 0.43 | -0.78 | 0.53 | 0.14 | 0.21 |
| eSOC SSST | 0.05 | 0.16 | 0.83 | 0.50 | -0.78 | 0.53 | 0.14 | 0.21 |
| WL SSST | 1.04 | 0.33 | 0.26 | 0.41 | 0.78 | 0.53 | 0.14 | 0.21 |

Notes: GO = goal-oriented intervention; WL = waiting list; SOC = standard of care; eSOC = enhanced standard of care; PE = psychoeducation; SSST = safe sex skills training; Edu = education.

##### *2.3 Design-by-treatment test*

chi2 (10) = 1.15
Prob > chi2 = 0.3769

***Appendix-16: Comparison-adjusted funnel plots***

*1. Proportion- proportion of condomless sex*

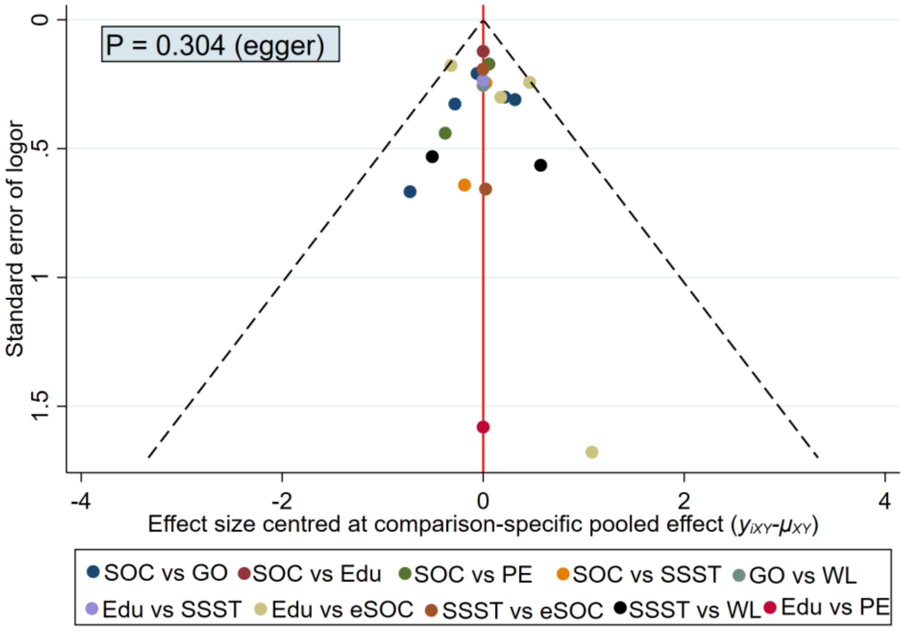

*2. Frequency- mean number of condomless sex in the past month*

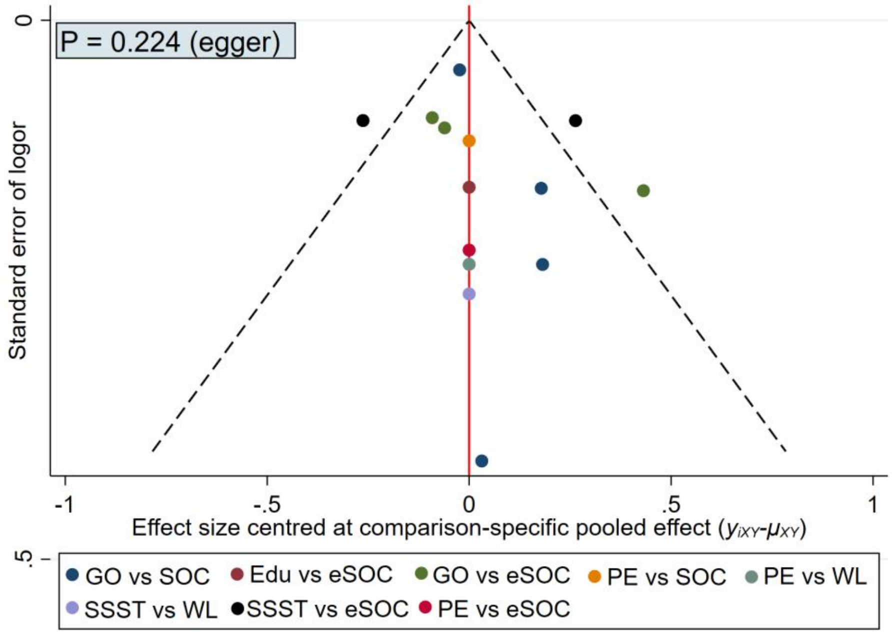

#### *Appendix-17: Sensitivity Analyses*

For each sensitivity analysis we present below:
- Network plot
- Results of statistical test for inconsistency of the network and common estimate for heterogeneity
- Forest-plot of results of the network-meta-analysis (reference TAU) - League-table of results of the network meta-analysis

#### *1.Excluding Studies that focused on newly infected HIV+ individuals*

##### ***1.1 Proportion of condomless sex ——exclude 2 studies***

1) Network plot

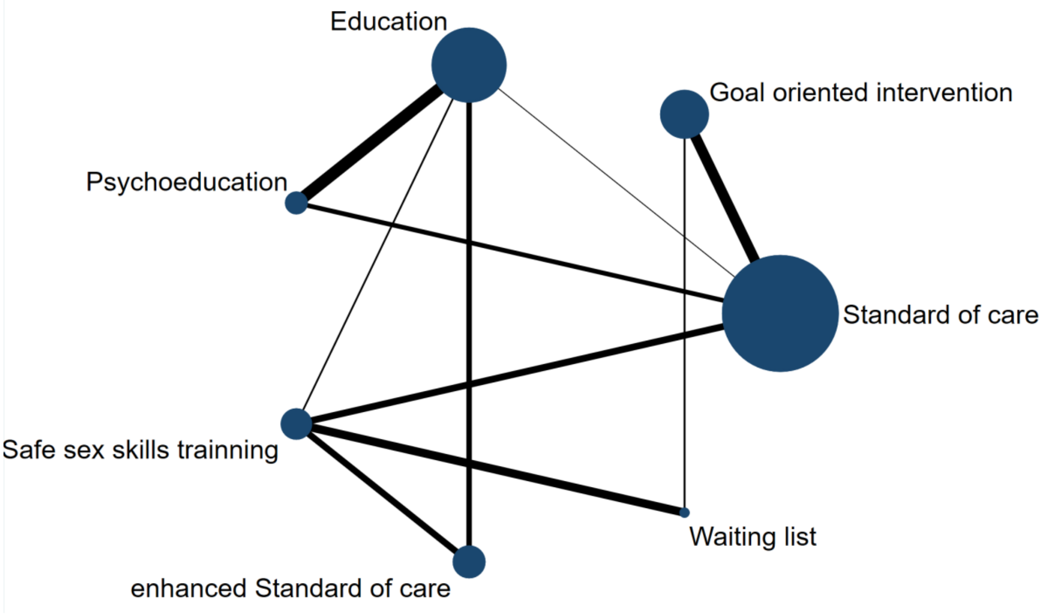

2) Inconsistency and heterogeneity

| Between study variance (tau) | P value of design-by-treatment interaction test | Inconsistent comparisons of detachable comparisons (%) (SIDE-test $p<0.05$) |
|---|---|---|
| 0.24 | 0.5326 | 0/7(0.00%) |

3) Forest-plot

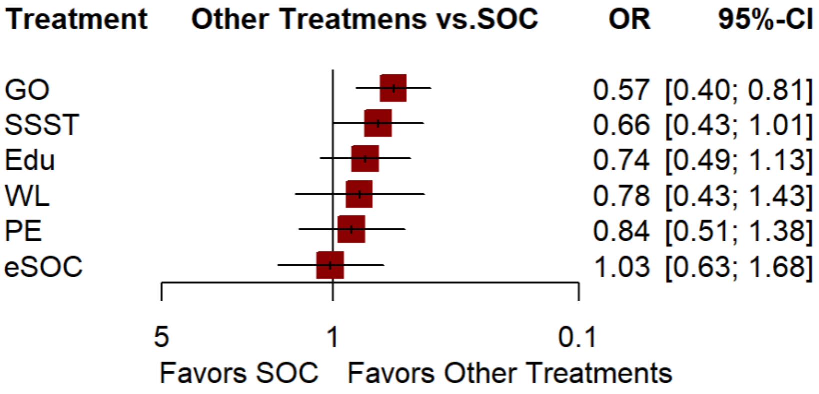

##### ***1.2 Mean number of condomless sex in the past month—— exclude 2 studies***

1) Network plot

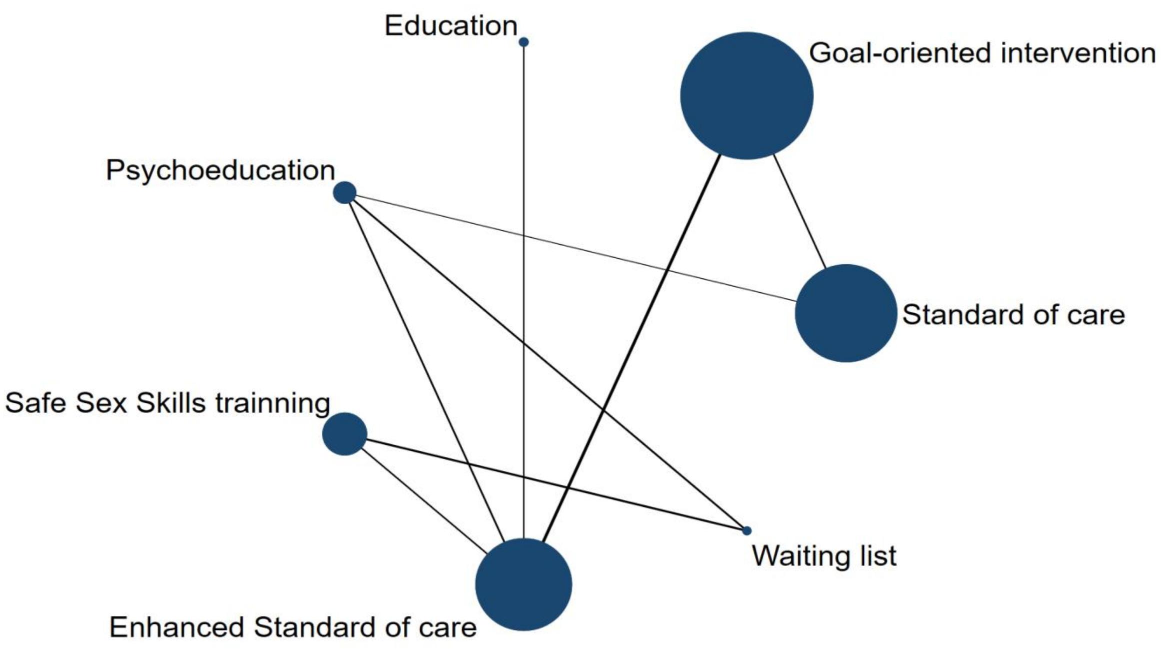

2) Inconsistency and heterogeneity

| Between study variance (tau) | P value of design-by-treatment interaction test | Inconsistent comparisons of detachable comparisons (%) (SIDE-test p＜0.05) |
|---|---|---|
| 0.25 | 0.4512 | 0/8(0.00%) |

3) Forest-plot

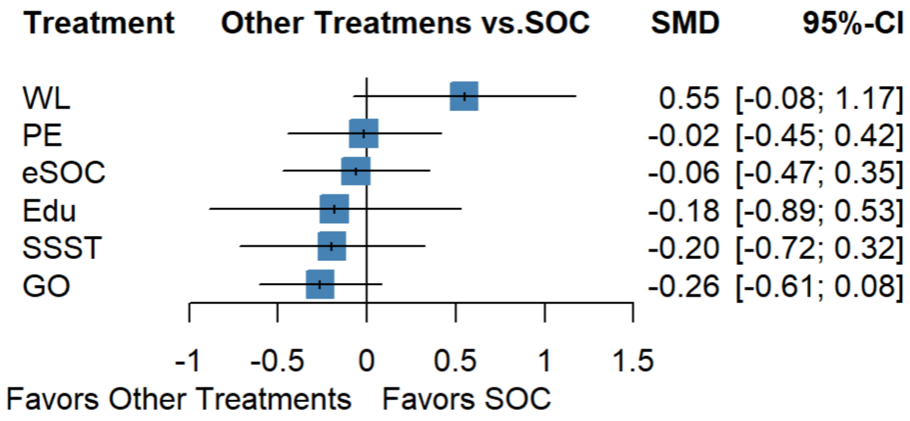

#### *2. Excluding Studies included HIV serodiscordant couples*

##### ***2.1 Proportion of condomless sex ——exclude 2 studies***

1) Network plot

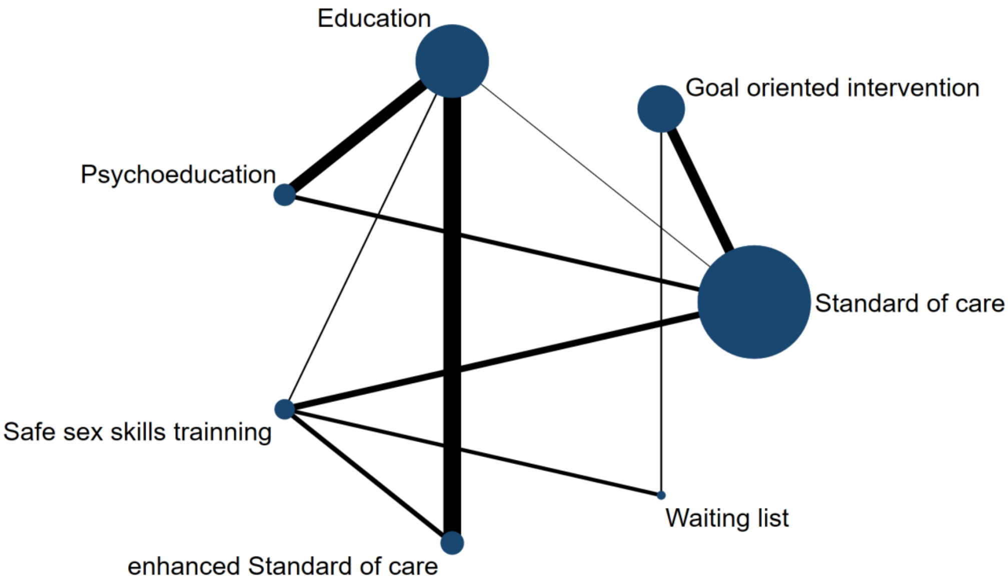

2) Inconsistency and heterogeneity

| Between study variance (tau) | P value of design-by-treatment interaction test | Inconsistent comparisons of detachable comparisons (%) (SIDE-test p＜0.05) |
|---|---|---|
| 0.19 | 0.6601 | 0/7(0.0%) |

3）Forest-plot

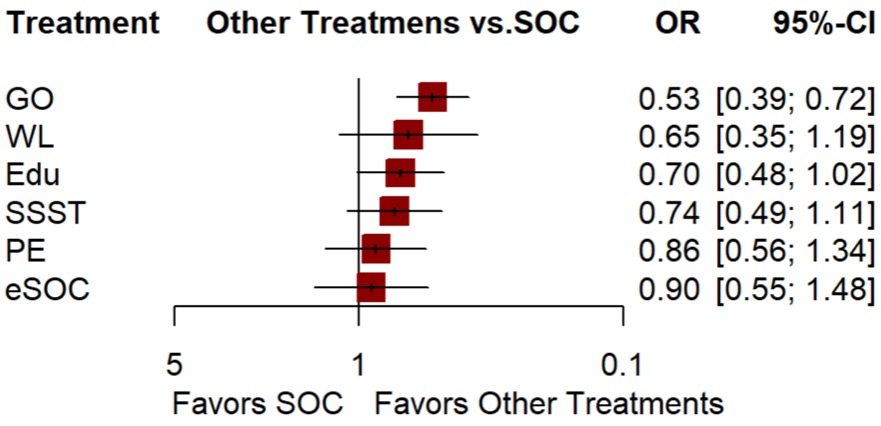

*2.2 Mean number of condomless sex in the past month——exclude 1 study*

1) Network plot

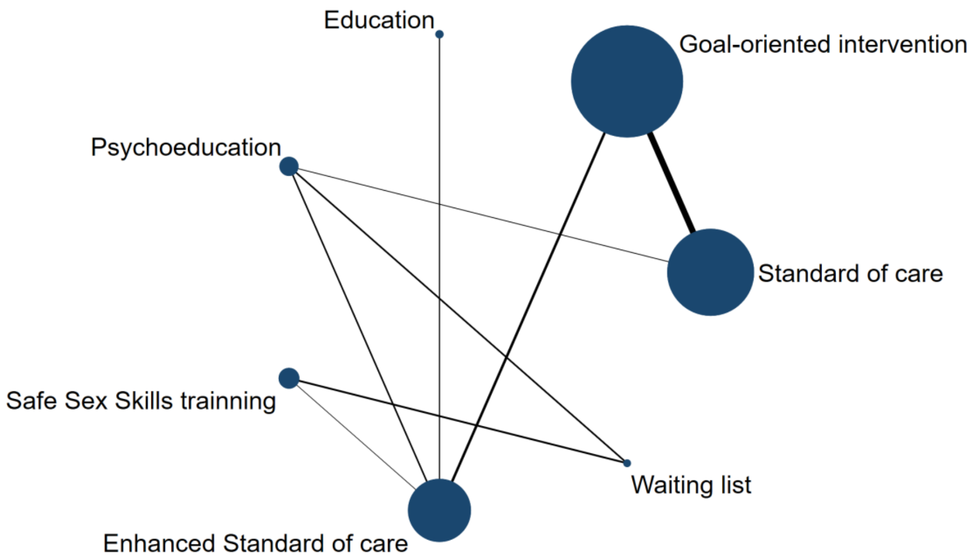

2) Inconsistency and heterogeneity

| Between study variance (tau) | P value of design-by-treatment interaction test | Inconsistent comparisons of detachable comparisons (%) (SIDE-test p＜0.05) |
|---|---|---|
| 0.21 | 0.1772 | 3/8(37.5%) |

3) Forest-plot

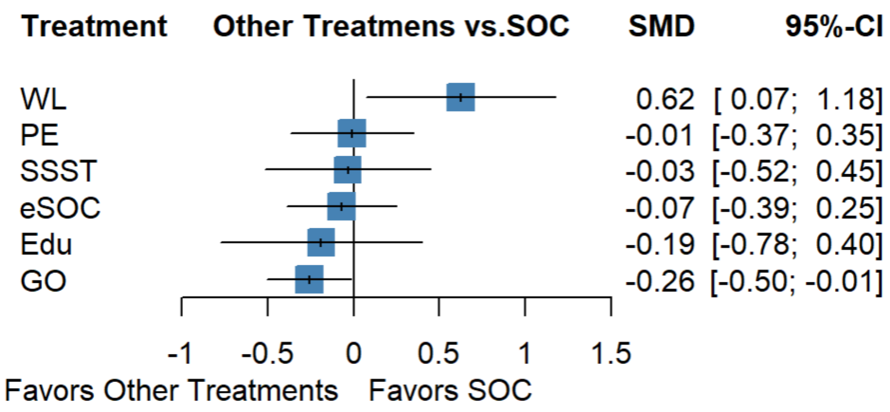

*3. Excluding Studies with Sample Sizes ≤ 20 in each group*

***3.1 Proportion of condomless sex ——exclude 3 studies***

1) Network plot

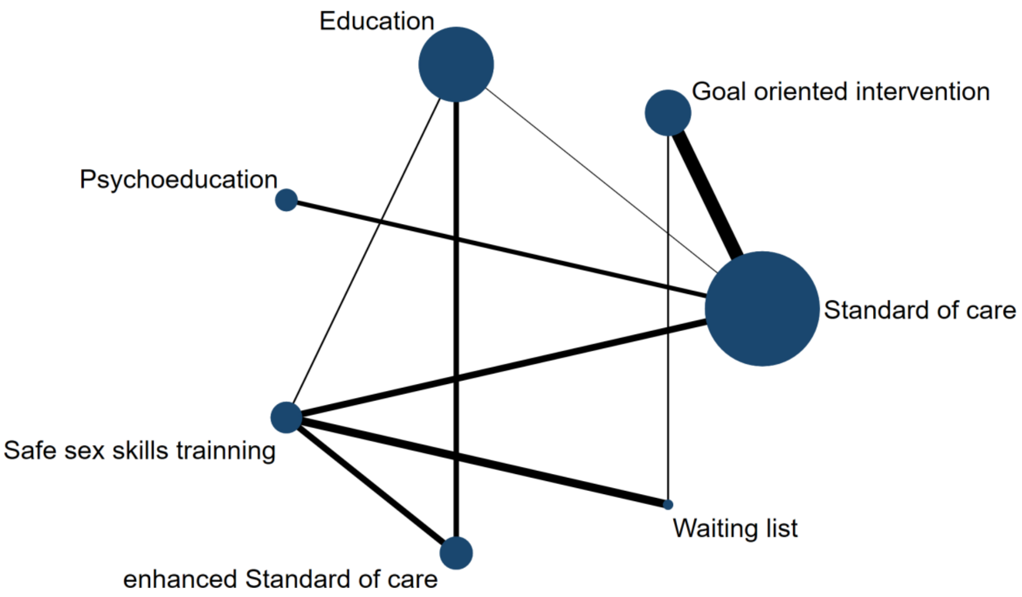

2) Inconsistency and heterogeneity

| Between study variance (tau) | P value of design-by-treatment interaction test | Inconsistent comparisons of detachable comparisons (%) (SIDE-test p＜0.05) |
|---|---|---|
| 0.21 | 0.4366 | 0/7(0.0%) |

3) Forest-plot

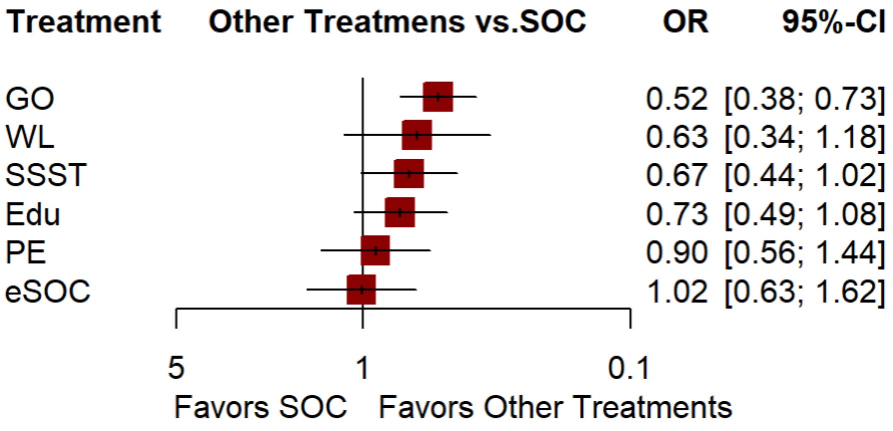

*3.2 Mean number of condomless sex in the past month——exclude 1 study*

1) Network plot

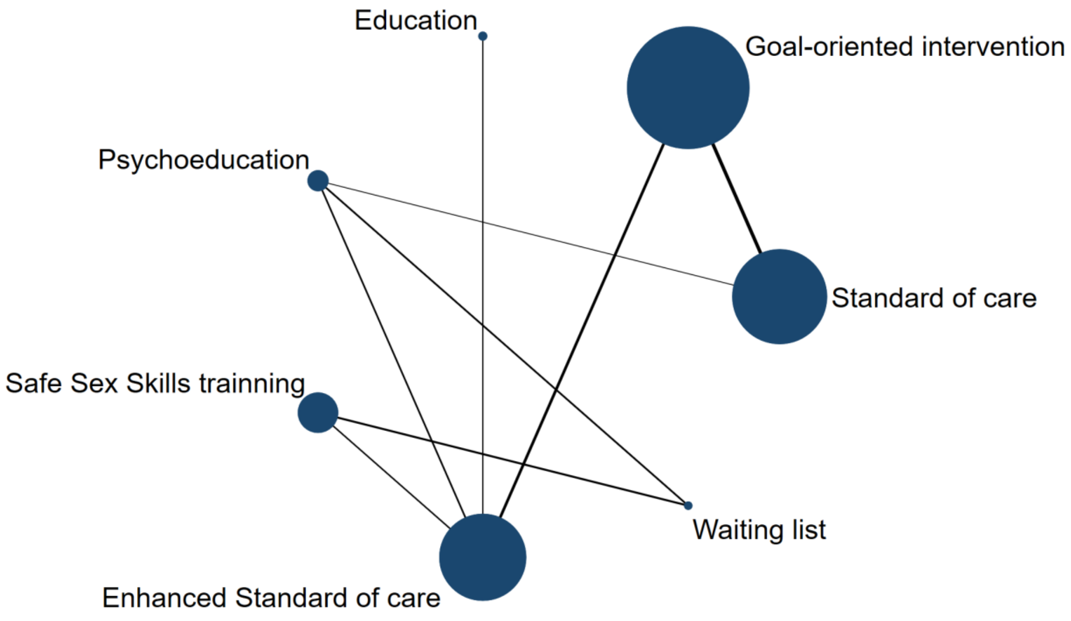

2) Inconsistency and heterogeneity

| Between study variance (tau) | P value of design-by-treatment interaction test | Inconsistent comparisons of detachable comparisons (%) (SIDE-test p＜0.05) |
|---|---|---|
| 0.23 | 0.4095 | 0/8(0.0%) |

3) Forest-plot

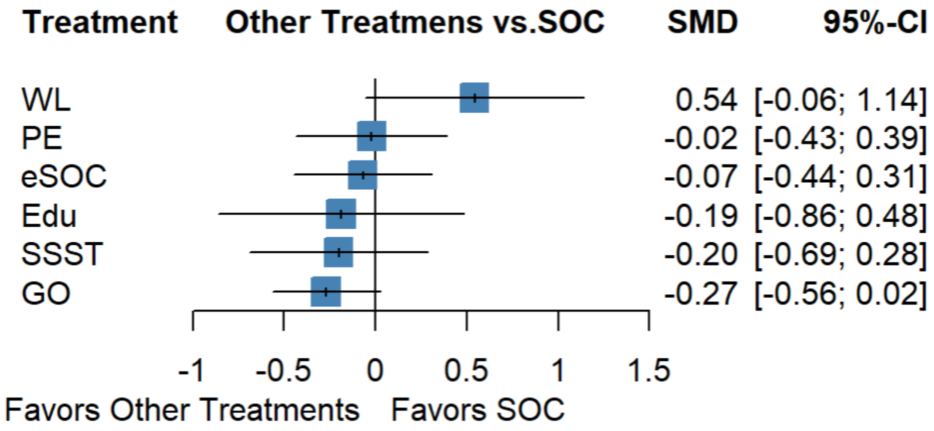

*4. Excluding studies rated as high risk of bias*

***4.1 Proportion of condomless sex ——exclude 8 studies***

1) Network plot

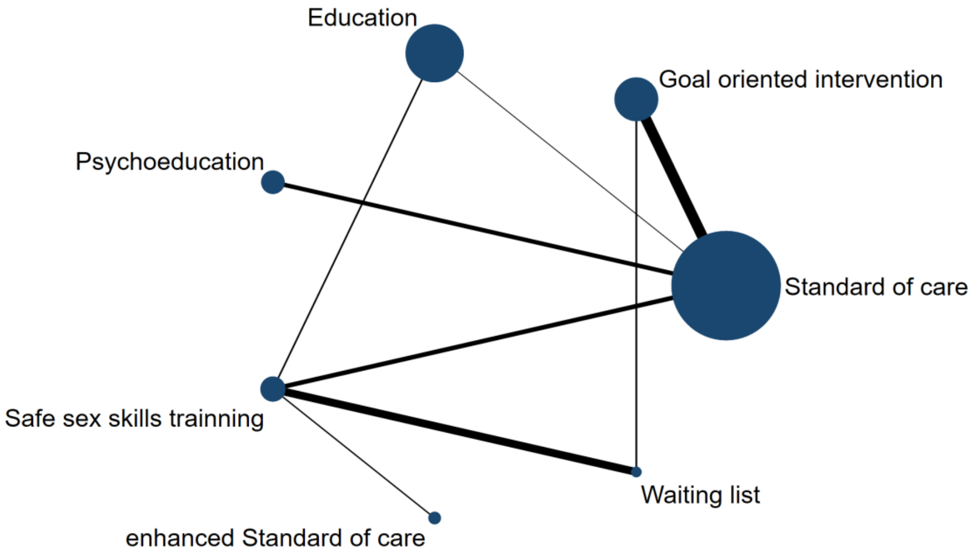

2) Inconsistency and heterogeneity

| Between study variance (tau) | P value of design-by-treatment interaction test | Inconsistent comparisons of detachable comparisons (%) (SIDE-test p＜0.05) |
|---|---|---|
| 0.24 | 0.8715 | 0/7(0%) |

3) Forest-plot

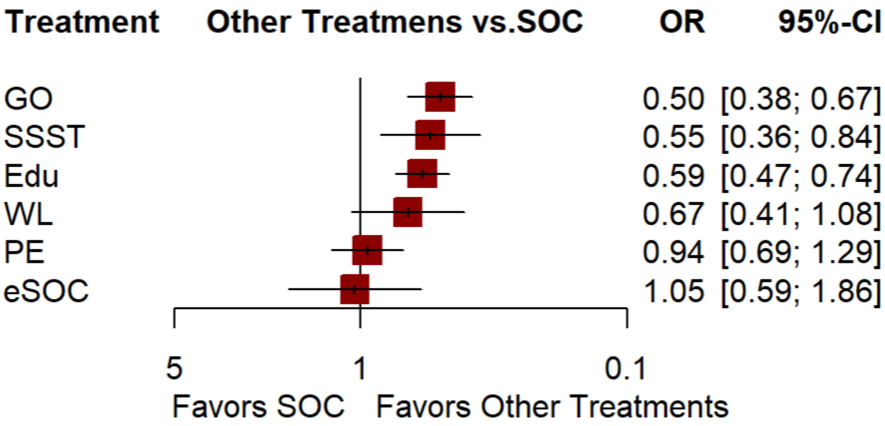

*4.2 Mean number of condomless sex in the past month——exclude 2 studies*

1) Network plot

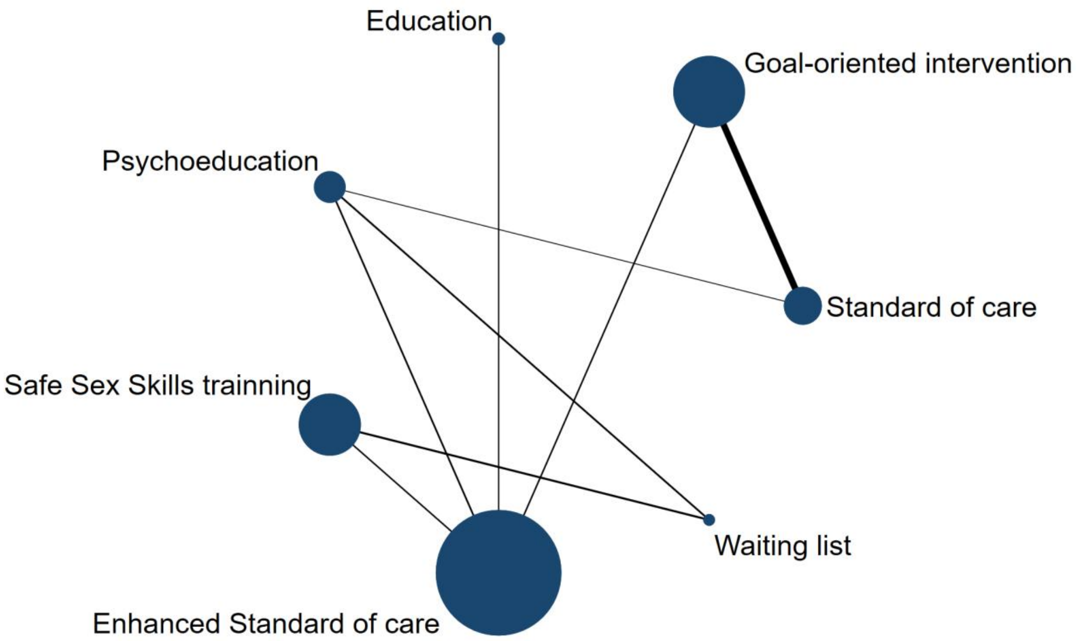

2) Inconsistency and heterogeneity

| Between study variance (tau) | P value of design-by-treatment interaction test | Inconsistent comparisons of detachable comparisons (%) (SIDE-test p＜0.05) |
|---|---|---|
| 0.19 | 0.4950 | 0/8(0.0%) |

3) Forest-plot

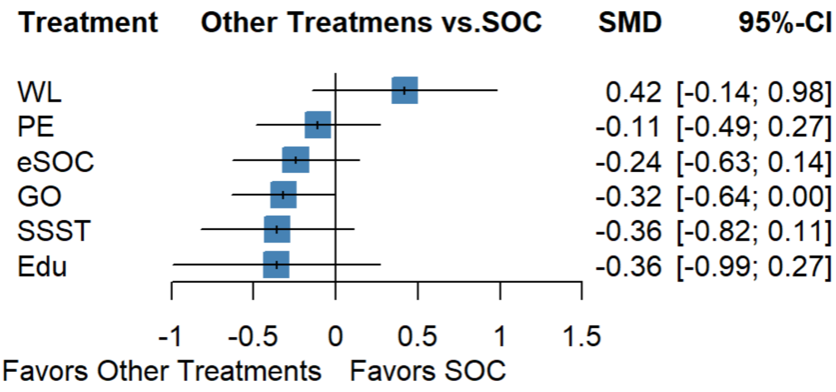

***Appendix-18: Summary of meta-analysis for sexual partner***

*1) Mean number of sexual partners*

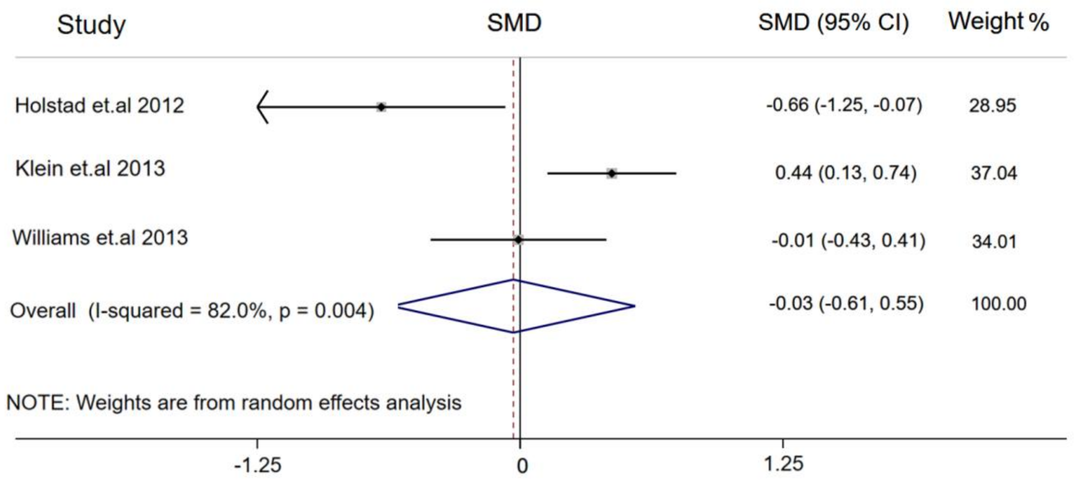

Publication of bias: egger test (p=0.009)
Estimate of between-study variance $Tau^2 = 0.2118$

*2) Proportion of multiple sexual partners*

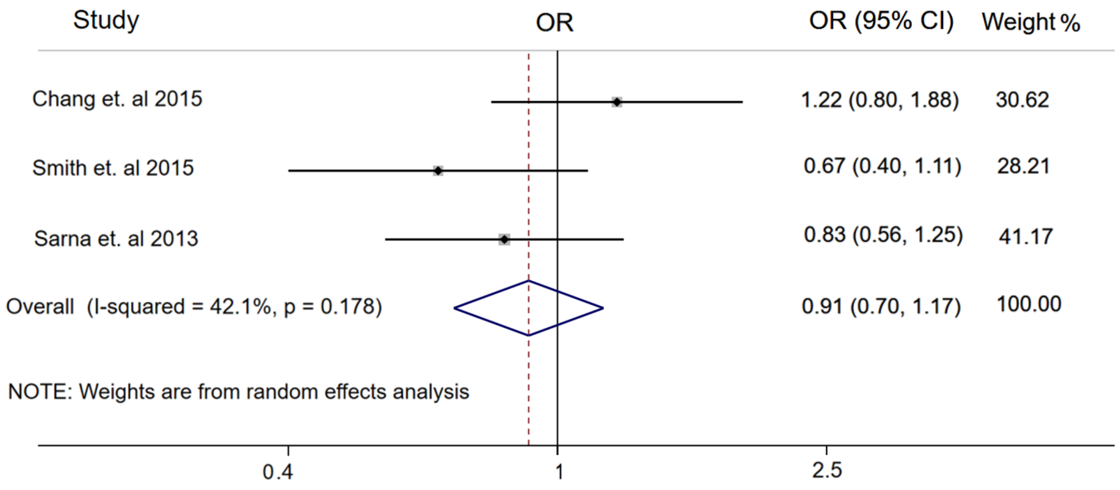

Publication of bias: egger test (p=0.679)
Estimate of between-study variance $Tau^2 = 0.0375$

#### *Appendix-19: GRADE Ratings for All Contrasts*

We accessed the confidence of this evidence based on the evaluation method for network meta-analyses developed by the GRADE Working Group. We assessed and graded the evidence based on seven domains, including within-study bias, indirectness, incoherence, inconsistency, heterogeneity, imprecision and reporting bias.

##### *1. Contribution of low, moderate, or high RoB comparisons to each network estimate*

The risk of bias of each study is reported in Appendix-5, We combined the results of ROB with the evidence contribution plot into two bar charts below, each bar shows how much information comes from comparisons at "low" [green], "some concerns" [yellow] or "high" [red] risk of bias.

*(1) Proportion- proportion of condomless sex*

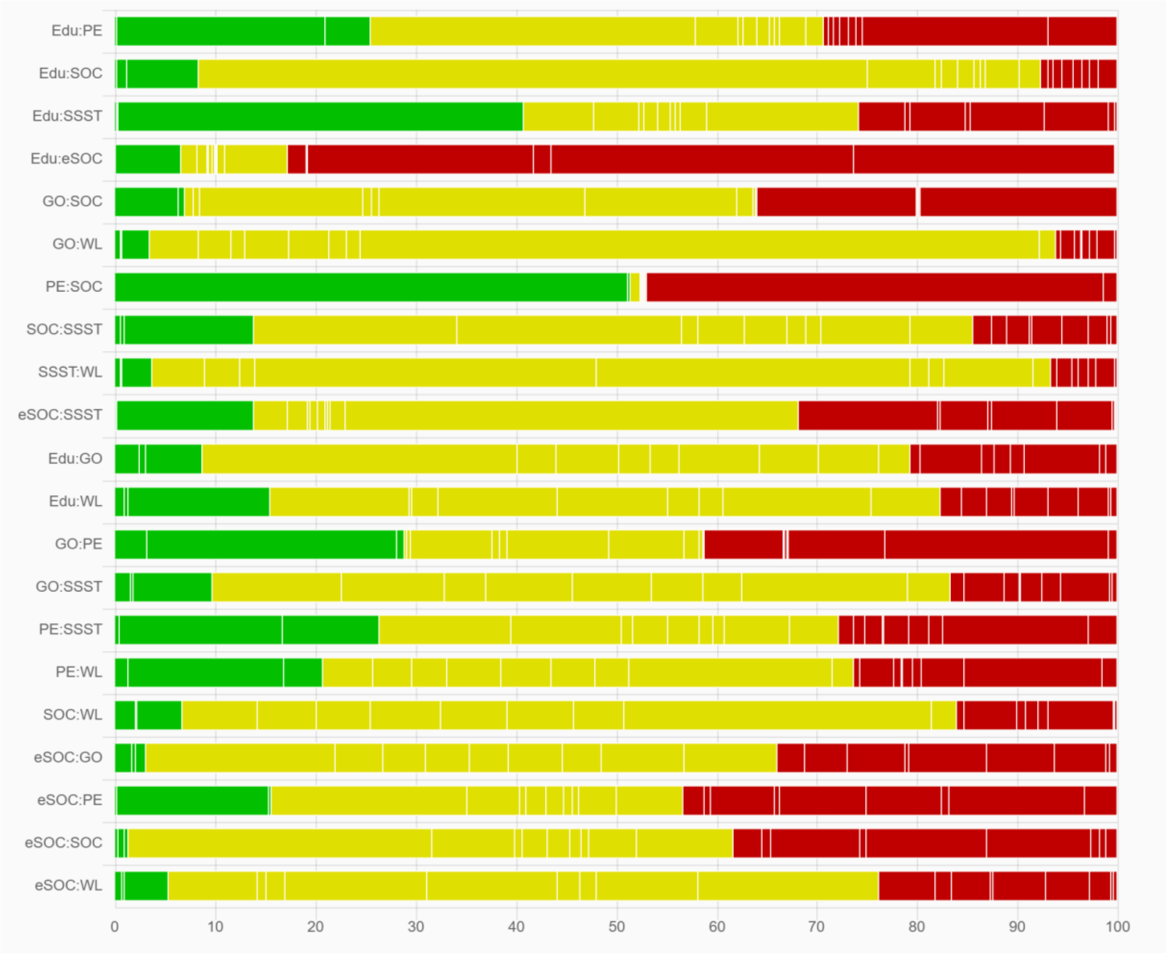

Notes: GO = goal-oriented intervention; WL = waiting list; SOC = standard of care; Edu = education; eSOC = enhanced standard of care; PE = psychoeducation; SSST = safe sex skills training.

*(2) Frequency- mean number of condomless sex in the past months*

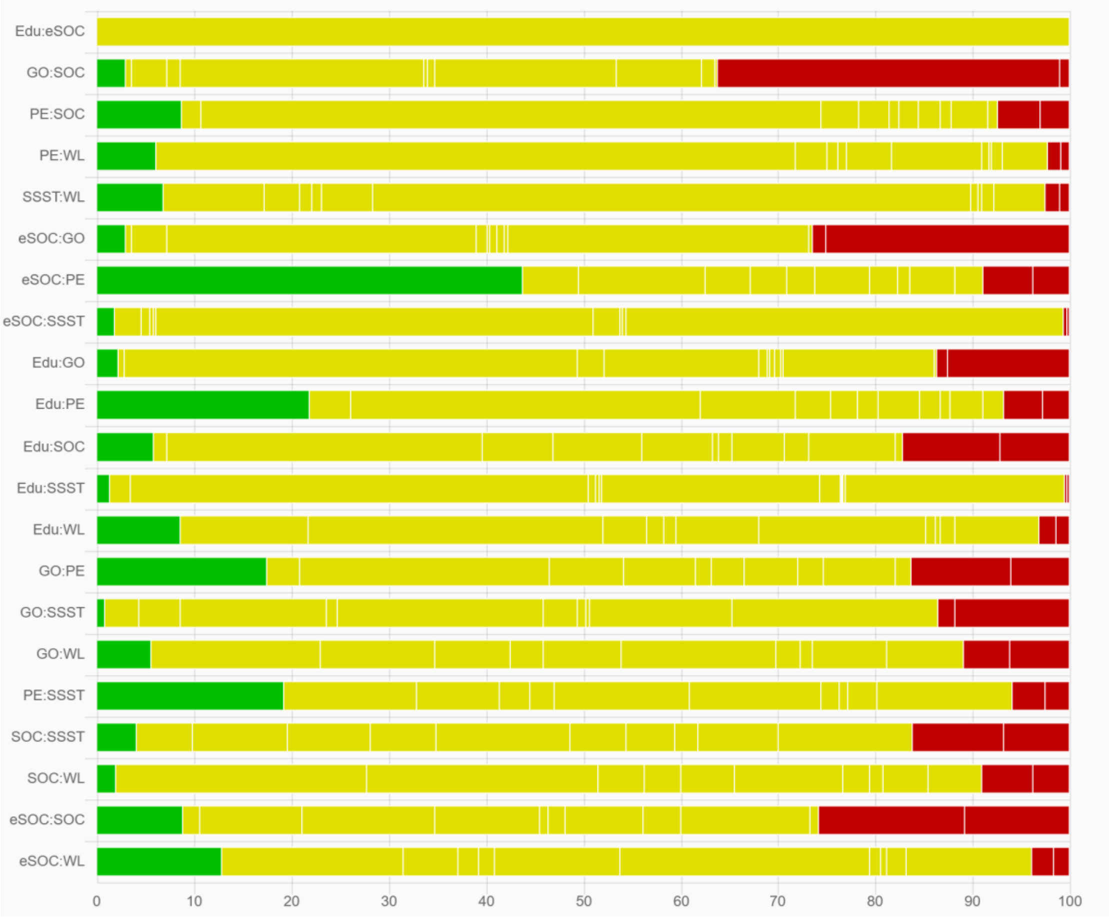

Notes: GO = goal-oriented intervention; WL = waiting list; SOC = standard of care; Edu = education; eSOC = enhanced standard of care; PE = psychoeducation; SSST = safe sex skills training.

#### *2. The results of GRADE Ratings for each comparison and ranking*

Based on all the above information, we used CINeMA, a recently developed web application based on the GRADE framework. Judgments for each domain can be summarized in CINeMA in an overall judgment on the confidence in the NMA estimate for each comparison. The level of confidence in the estimate can be classified as 'very low', 'low', 'moderate' and 'high'.

The meaning of these four levels can be interpreted as follows:

High quality: Further research is very unlikely to change our confidence in the estimate of effect.

Moderate quality: Further research is likely to have an important impact on our confidence in the estimate of effect and may change the estimate.

Low quality: Further research is very likely to have an important impact on our confidence in the estimate of effect and is likely to change the estimate.

Very low quality: We are very uncertain about the estimate.

The CINeMA-guidance-document suggests for each comparison to start at the first level (i.e. high) and to downgrade for one level for a rating of "some concerns", and by two levels for a rating of "major concerns"[17]. It is recommended to consider judgements on different domains jointly rather than in isolation. The reason is that domains are interconnected and downgrading more than once for related concerns should be avoided. Based on these recommendations, we used the following approach to reach an overall level of confidence for each comparison and outcome:

1 judgement of "some concerns" leads to downgrading by 1 level (e.g. from 'high' to 'moderate');

1 judgement of "major concerns" leads to downgrading by 2 levels (e.g. from 'high' to 'low');

2 judgements of "some concerns" could be interconnected and do not justify downgrading more than by 1 level;

1 judgement of "major concerns" and up to 2 judgements of "some concerns" or 1 additional judgement of "major concerns" could be interconnected and do not justify downgrading by more than 2 levels;

2 judgements of "major concerns" and any additional judgements of "some concerns" or "major concerns" (or more than 4 judgements of some concerns) lead to downgrading by three levels (the maximum).

*(1) Proportion of condomless sex*

| Comparison | Number of Studies | Within-study bias | Reporting bias | Indirectness | Imprecision | Heterogeneity | Incoherence | Confidence rating |
|---|---|---|---|---|---|---|---|---|
| Mixed evidence | | | | | | | | |
| Edu vs PE | 1 | Some concerns | Low risk | No concerns | Some concerns | Some concerns | No concerns | Moderate |
| Edu vs SOC | 1 | Some concerns | Low risk | No concerns | No concerns | Some concerns | No concerns | Moderate |
| Edu vs SSST | 1 | No concerns | Low risk | No concerns | Some concerns | Some concerns | No concerns | Moderate |
| Edu vs eSOC | 4 | Major concerns | Low risk | No concerns | Some concerns | Some concerns | No concerns | Very low |
| GO vs SOC | 6 | Some concerns | Low risk | No concerns | No concerns | No concerns | No concerns | Moderate |
| GO vs WL | 1 | Some concerns | Low risk | No concerns | Major concerns | No concerns | No concerns | Low |
| PE vs SOC | 2 | No concerns | Low risk | No concerns | Major concerns | No concerns | No concerns | Low |
| SOC vs SSST | 1 | Some concerns | Low risk | No concerns | No concerns | Some concerns | No concerns | Moderate |
| SSST vs WL | 2 | Some concerns | Low risk | No concerns | Major concerns | No concerns | No concerns | Low |
| SSST vs eSOC | 2 | Some concerns | Low risk | No concerns | No concerns | Some concerns | No concerns | Moderate |
| Indirect evidence | | | | | | | | |
| Edu vs GO | -- | Some concerns | Low risk | No concerns | Major concerns | No concerns | No concerns | Low |
| Edu vs WL | -- | Some concerns | Low risk | No concerns | Major concerns | No concerns | No concerns | Low |

| | | | | | | | | |
|---|---|---|---|---|---|---|---|---|
| GO vs PE | -- | Some concerns | Low risk | No concerns | Some concerns | No concerns | No concerns | Moderate |
| GO vs SSST | -- | Some concerns | Low risk | No concerns | Major concerns | No concerns | No concerns | Low |
| GO vs WL | -- | Some concerns | Low risk | No concerns | Major concerns | No concerns | No concerns | Low |
| GO vs eSOC | -- | Some concerns | Low risk | No concerns | Some concerns | Some concerns | No concerns | Moderate |
| PE vs SSST | -- | Some concerns | Low risk | No concerns | Some concerns | Some concerns | No concerns | Moderate |
| PE vs WL | -- | Some concerns | Low risk | No concerns | Major concerns | No concerns | No concerns | Low |
| PE vs eSOC | -- | Some concerns | Low risk | No concerns | Major concerns | No concerns | No concerns | Low |
| SOC vs WL | -- | Some concerns | Low risk | No concerns | Major concerns | No concerns | No concerns | Low |
| SOC vs eSOC | -- | Some concerns | Low risk | No concerns | Major concerns | No concerns | No concerns | Low |
| WL vs eSOC | -- | Some concerns | Low risk | No concerns | Major concerns | No concerns | No concerns | Low |

Notes: GO = goal-oriented intervention; WL = waiting list; SOC = standard of care; Edu = education; eSOC = enhanced standard of care; PE = psychoeducation; SSST = safe sex skills training.

*(2) Mean number of condomless sex in the past months*

| Comparison | Number of Studies | Within-study bias | Reporting bias | Indirectness | Imprecision | Heterogeneity | Incoherence | Confidence rating |
|---|---|---|---|---|---|---|---|---|
| Mixed evidence | | | | | | | | |
| Edu vs eSOC | 1 | Some concerns | Low risk | No concerns | Major concerns | No concerns | No concerns | Low |
| GO vs SOC | 4 | Some concerns | Low risk | No concerns | Some concerns | Some concerns | No concerns | Moderate |
| GO vs eSOC | 3 | Some concerns | Low risk | No concerns | Some concerns | Some concerns | No concerns | Moderate |
| PE vs SOC | 1 | Some concerns | Low risk | No concerns | Major concerns | No concerns | No concerns | Low |
| PE vs WL | 1 | Some concerns | Low risk | No concerns | No concerns | Major concerns | No concerns | Low |
| PE vs eSOC | 1 | Some concerns | Low risk | No concerns | Major concerns | No concerns | No concerns | Low |
| SSST vs WL | 1 | Some concerns | Low risk | No concerns | No concerns | Some concerns | No concerns | Moderate |
| SSST vs eSOC | 2 | Some concerns | Low risk | No concerns | Some concerns | Some concerns | No concerns | Moderate |
| Indirect evidence | | | | | | | | |
| Edu vs GO | -- | Some concerns | Low risk | No concerns | Major concerns | No concerns | No concerns | Low |
| Edu vs PE | -- | Some concerns | Low risk | No concerns | Major concerns | No concerns | No concerns | Low |
| Edu vs SOC | -- | Some concerns | Low risk | No concerns | Major concerns | No concerns | No concerns | Low |
| Edu vs SSST | -- | Some concerns | Low risk | No concerns | Major concerns | No concerns | No concerns | Low |
| Edu vs WL | -- | Some concerns | Low risk | No concerns | Some concerns | Some concerns | No concerns | Moderate |

| | | | | | | | | |
|---|---|---|---|---|---|---|---|---|
| GO vs PE | -- | Some concerns | Low risk | No concerns | Some concerns | Some concerns | No concerns | Moderate |
| GO vs SSST | -- | Some concerns | Low risk | No concerns | Major concerns | No concerns | No concerns | Low |
| GO vs WL | -- | Some concerns | Low risk | No concerns | No concerns | Some concerns | No concerns | Moderate |
| PE vs SSST | -- | Some concerns | Low risk | No concerns | Major concerns | No concerns | No concerns | Low |
| SOC vs SSST | -- | Some concerns | Low risk | No concerns | Major concerns | No concerns | No concerns | Moderate |
| SOC vs WL | -- | Some concerns | Low risk | No concerns | Some concerns | Some concerns | No concerns | Moderate |
| SOC vs eSOC | -- | Some concerns | Low risk | No concerns | Major concerns | No concerns | No concerns | Low |
| WL vs eSOC | -- | Some concerns | Low risk | No concerns | No concerns | Major concerns | No concerns | Low |

Notes: GO = goal-oriented intervention; WL = waiting list; SOC = standard of care; Edu = education; eSOC = enhanced standard of care; PE = psychoeducation; SSST = safe sex skills training.

#### *References:*

1. Becasen JS, Denard CL, Mullins MM, Higa DH, Sipe TA. Estimating the Prevalence of HIV and Sexual Behaviors Among the US Transgender Population: A Systematic Review and Meta-Analysis, 2006-2017. *Am J Public Health* 2019; **109**(1): e1-e8.
2. Zhang H, Yu Q, Li Z, et al. Efficacy of Psychological Interventions Towards the Reduction of High-Risk Sexual Behaviors Among People Living with HIV: A Systematic Review and Meta-analysis, 2010-2020. *AIDS Behav* 2021; **25**(10): 3355-76.
3. Gross A, Schirm S, Scholz M. Ycasd - a tool for capturing and scaling data from graphical representations. *BMC Bioinformatics* 2014; **15**: 219.
4. Napper LE, Fisher DG, Reynolds GL, Johnson ME. HIV risk behavior self-report reliability at different recall periods. *AIDS and Behavior* 2010; **14**: 152-61.
5. Kanters S, Park JJ, Chan K, et al. Interventions to improve adherence to antiretroviral therapy: a systematic review and network meta-analysis. *The lancet HIV* 2017; **4**(1): e31-e40.
6. APA Dictionary of Psychology.  American Psychological Association Dictionary of Psychology ： wait-list control group.
7. Mann T, de Ridder D, Fujita K. Self-regulation of health behavior: social psychological approaches to goal setting and goal striving. *Health Psychol* 2013; **32**(5): 487-98.
8. Pearson ES. Goal setting as a health behavior change strategy in overweight and obese adults: a systematic literature review examining intervention components. *Patient Educ Couns* 2012; **87**(1): 32-42.
9. Bighelli I, Rodolico A, García-Mieres H, et al. Psychosocial and psychological interventions for relapse prevention in schizophrenia: a systematic review and network meta-analysis. *Lancet Psychiatry* 2021; **8**(11): 969-80.
10. Yu Y, Wang X, Wu Y, et al. The benefits of psychosocial interventions for mental health in men who have sex with men living with HIV: a systematic review and meta-analysis. *BMC Psychiatry* 2022; **22**(1): 440.
11. Turner RM, Jackson D, Wei Y, Thompson SG, Higgins JP. Predictive distributions for between‐study heterogeneity and simple methods for their application in Bayesian meta‐analysis. *Statistics in medicine* 2015; **34**(6): 984-98.
12. Rhodes KM, Turner RM, Higgins JP. Predictive distributions were developed for the extent of heterogeneity in meta-analyses of continuous outcome data. *Journal of clinical epidemiology* 2015; **68**(1): 52-60.
13. Andrade C. Mean Difference, Standardized Mean Difference (SMD), and Their Use in Meta-Analysis: As Simple as It Gets. *J Clin Psychiatry* 2020; **81**(5).
14. Zhao T, Tang C, Yan H, Lu Q, Guo M, Wang H. Comparative efficacy and acceptability of non-pharmacological interventions for depression in people living with HIV: A systematic review and network meta-analysis. *Int J Nurs Stud* 2023; **140**: 104452.
15. Brignardello-Petersen R, Murad MH, Walter SD, et al. GRADE approach to rate the certainty from a network meta-analysis: avoiding spurious judgments of imprecision in sparse networks. *J Clin Epidemiol* 2019; **105**: 60-7.
16. Brignardello-Petersen R, Bonner A, Alexander PE, et al. Advances in the GRADE approach to rate the certainty in estimates from a network meta-analysis. *J Clin Epidemiol* 2018; **93**: 36-44.
17. *Nikolakopoulou, A., Higgins, J. P., Papakonstantinou, T., Chaimani, A., Del Giovane, C., Egger, M., & Salanti, G. (2020). CINeMA: an approach for assessing confidence in the results of a network meta-analysis. PLoS medicine, 17(4), e1003082.*
